# Spatial analyses of infection and disease during chikungunya and Zika epidemics in Managua, Nicaragua

**DOI:** 10.64898/2026.09.15.26363072

**Authors:** Fausto Andres Bustos Carrillo, Brenda Lopez Mercado, Jairo Carey Monterrey, Damaris Collado, Tatiana Miranda, Saira Saborio, Sergio Ojeda, Nery Sanchez, Miguel Plazaola, Harold Suazo Laguna, Sonia Arguello, Hugh Sturrock, Angel Balmaseda, Guillermina Kuan, Eva Harris

## Abstract

Chikungunya (CHIKV) and Zika (ZIKV) viruses have caused widespread epidemics across the Americas. After several years of minimal transmission, CHIKV is once again spreading in Nicaragua. We investigated spatial aspects of two chikungunya epidemics (2014, 2015) and one Zika epidemic (2016) in a prospective pediatric cohort in Nicaragua. We used generalized estimating equations, generalized additive and mixed-effects models, Kuldorff’s scan statistic, and intracluster correlations to analyze the infection and disease status of ∼3,000 initially uninfected participants during each epidemic. We found that the incidence of infection (# new infections/total population) and disease (# new cases/total population) often had different spatial patterns, demonstrating that high-infection areas may not exhibit much disease. High infection incidence was observed near a large cemetery west of the study site. The large 2015 chikungunya epidemic and 2016 Zika epidemic started near the cemetery, exhibited high infection incidence throughout the study site, and had extensive month-to-month changes in spatiotemporal dynamics. Clusters of excess infections were large and adjacent to the cemetery, while clusters of excess uninfected participants were found in the northern and eastern sections of the study area. Notably, the intracluster correlation of infection was very weak in households (∼0.2), and infection status was correlated across distances <200 meters. Similarities across the epidemics suggest that targeting interventions to the built, non-household environment may reduce epidemic potential in Managua. Overall, spatial analyses of epidemics that only use case data may substantially underestimate the full extent of viral transmission, missing identification of high-infection areas and opportunities for intervention.

## INTRODUCTION

Chikungunya virus (CHIKV) [1] and Zika virus (ZIKV) [1] are pathogens of significant human concern that are transmitted by *Aedes aegypti* and *Aedes albopictus* mosquitoes. CHIKV and ZIKV have caused massive epidemics since their introduction to the Americas in 2013 and 2015, respectively [1]. As of mid-August 2026, over 6.13 million cases (symptomatic infections) of chikungunya and 1.06 million cases of Zika in the Americas have been reported to the Pan American Health Organization [2]. However, the true number of cases is likely much higher, as the diseases can present with mild manifestations, particularly in children, that lead to misdiagnoses [3]. Further, the widespread use of febrile surveillance systems to capture cases may well lead to under-diagnosis since Zika often manifests without fever [4]. Acute chikungunya typically presents with high fever and crippling, transient arthralgia. Chronic chikungunya manifests with arthralgia that can persist for months or years after acute illness [1]. Zika typically presents non-specifically during childhood and as a mild, dengue-like illness during adolescence and adulthood [4]. ZIKV infection may infrequently trigger Guillain-Barré Syndrome in adults [1]; infection during pregnancy can cause an array of neurodevelopmental complications in the infant, called Congenital Zika Syndrome, including microcephaly [1].

For dengue virus, which is spread by the same mosquitoes in similar areas around the world as CHIKV and ZIKV, households are generally thought to be key transmission sites [5,6], although the evidence is mixed [7,8]. In Bangladesh, households have been identified as important sites for CHIKV transmission [9], but similar evidence from the Americas is limited. To document the spatial characteristics of transmission, studies of infectious diseases typically obtain data on cases retroactively using febrile surveillance systems, as the routine, large-scale measurement of infection status is difficult and expensive. Since infections by ZIKV as well as the Asian lineage of CHIKV (the first CHIKV lineage to circulate in Central America) are often clinically inapparent (asymptomatic) [1,10,11], case-only data cannot fully characterize epidemics of these viruses.

The Pediatric Dengue Cohort Study (PDCS), located in District II of Managua, Nicaragua, was initiated in 2004 to study dengue virus and later extended to include CHIKV and ZIKV [3]. The PDCS investigates arthropod-borne viruses and is the longest continuously running cohort study in the arboviral field. With its prospective study design and participant households’ geolocation data, the PDCS is well situated to estimate both the infection incidence (infections / total population) and disease incidence (cases / total population) across its study area and over epidemic seasons. Although Zika case counts in Nicaragua are currently at low levels, chikungunya case counts are increasing sharply as a new epidemic initiates. To serve as a benchmark for future studies, we examined historical data from the two large chikungunya epidemics in the cohort in 2014 and 2015 [12,13], termed ChikE1 and ChikE2, respectively. For comparison, we also examined the explosive 2016 Zika epidemic [4,12], ZikaE, which is the largest epidemic ever experienced by the PDCS. Here, we use epidemic-specific datasets and point data to present spatial and spatiotemporal analyses. This study maps the incidence of infection and disease across the study area, tests whether infections cluster within households, identifies geographic clusters of infection and disease, estimates risk factors for infection and disease after controlling for spatial correlation, and characterizes the spatiotemporal dynamics of infection and disease occurrence.

## MATERIALS AND METHODS

### Ethics statement

The Pediatric Dengue Cohort Study (PDCS) was approved by Institutional Review Boards (IRBs) of the University of California, Berkeley; the University of Michigan, Ann Arbor; and the Nicaraguan Ministry of Health. Adult participants and the parents/legal guardians of pediatric participants provided written informed consent. Subjects six years and older provided verbal assent. This study was performed in accordance with the principles stated in the Declaration of Helsinki.

### Study design

The PDCS [13] is an open, population-based, prospective cohort of Nicaraguan children. We assessed ∼3,000 PDCS participants 2-14 years old who experienced two chikungunya epidemics and one Zika epidemic from September 2014 to January 2017. Analysis of each epidemic was restricted to participants who lived within the health center’s catchment area (Fig. S1), were enrolled before each epidemic began, and were immunologically naïve to the incoming virus at the start of each epidemic.

The catchment area of the study health center, the Health Center Sócrates Flores Visas (HCSFV), consists of 18 neighborhoods; however, no PDCS participants resided in the smallest neighborhood, Mantica, during the study period. The population served by the HCSFV is approximately 62,000 persons [14]. The catchment area of the HCSFV and hence the study area is approximately 5 km^2^ and is approximately 3 km long at its widest. The 18 neighborhoods that constitute the study area reside in District II of Managua, Nicaragua. The study area is low-lying (42-106 meters above sea level), located below Lago Xolotlán (Lake Managua), and quite flat. About 88% of PDCS households have garbage collection services and about 95% have sewage systems and tap water [14], although tap water may not be potable or available for all hours of the day.

As previously published [13], participants were initially recruited into the PDCS during door-to-door visits of households served by the HCSFV in 2004, during which eligible children were invited to participate. In addition to parental and participant consent, the eligibility criteria for the PDCS included the following: being between 2 and 9 years old, living in the catchment area of the HCSFV, having no plans to leave the catchment area within a period of 3 years, attending the HCSFV for all medical needs, and lacking any immunocompromising medical conditions. Since 2004, the PDCS has been expanded to include children between the ages of 2 and 14. Recruitment into the PDCS occurs every year to ensure the age structure remains constant.

### Inclusion and exclusion criteria

All PDCS participants for whom infection status could be determined and who were immunologically naïve to CHIKV or ZIKV at the beginning of beginning of the chikungunya and Zika epidemics were initially considered eligible to participate in this study. Enrolled participants who were then identified as violating the eligibility criteria of the PDCS, or participants for whom spatial analyses could not be conducted (*e.g.*, due to missing global positioning system (GPS) points of their household), were excluded from this study. In particular, we excluded participants from this analysis who were enrolled after the start of a given epidemic. Although the infection status of such participants could be serologically determined after the epidemic (with paired pre- and post-epidemic annual samples), they were not under disease surveillance between the start of the epidemic and their enrollment date. As a result, participants who were enrolled after an epidemic started and who had an acute illness of either chikungunya or Zika before their enrollment date would have been incorrectly recorded as having experienced clinically inapparent infections. The bias toward inapparent infections among this set of participants led us to exclude them. This criterion was the major reason for the exclusion of participants; it also led to the analysis of a closed cohort of initially uninfected participants who subsequently experienced an epidemic, even though the PDCS is an open cohort study.

For example, there were 3,808 enrolled PDCS participants for whom ZIKV infection data, as measured by the NS1 blockade-of-binding enzyme-linked immunosorbent assay, were available for analysis of the Zika epidemic (ZikaE). Of those, 39 were dropped because they were not under disease surveillance during any point between January 1, 2016 and January 31, 2017 (a time period that covers the full duration of the Zika epidemic [4]). Ten participants were dropped for lack of GPS points, which were necessary for all spatial analyses. An additional 22 individuals were dropped for having moved outside the catchment area of the HCSFV since enrollment into the study [13]. Of the remaining 3,737 participants, 720 participants were dropped for being enrolled after the Zika epidemic started, resulting in a total of 3,017 PDCS participants who were at risk of ZIKV infection at the beginning of the epidemic, under disease surveillance at that time, met PDCS eligibility criteria, and had GPS data that were available to analyze for the present study.

The application of these criteria resulted in a similar number of exclusions for PDCS participants during the first chikungunya epidemic (ChikE1) and the second chikungunya epidemic (ChikE2). The immunologically naïve population for ChikE2 (n=2,864) is the smallest we considered because all eligible PDCS participants who were infected during ChikE1 were no longer at risk of incident CHIKV infection during ChikE2. These participants were consequently excluded from analysis of ChikE2. Between the beginning of the 2016 annual serosurvey and the end of the 2017 annual serosurvey, we detected five new CHIKV infections. However, because these five CHIKV infections that occurred in 2016 numbered too few for spatial analyses, they were also excluded from the present study.

### Participants’ annual serosurveys, questionnaires, and primary care

Between March and April of each year, healthy PDCS participants visit the HCSFV to provide a blood sample [13,15]. During this annual serosurvey, questionnaires are administered orally to PDCS participants and their family members. Age, sex, level of education, and other demographic/anthropometric information is collected on participant questionnaires. A household questionnaire is used to collect information on assets and conditions of the household.

As part of the PDCS protocol, families agree to bring study participants to the HCSFV, where they receive free primary care 24/7, at the first indication of any illness. PDCS participants with undifferentiated febrile illness or with suspected chikungunya or Zika provide acute and convalescent blood samples during clinical visits [4]. During the last satisfaction survey we conducted of the entire cohort, 96% of participants rated the medical attention received at the HCSFV as either excellent or very good [13]. In participant surveys, an average of 2% of participants reported attending a health care provider other than the HCSFV, and an average of 3% of participants reported not seeking any medical attention during an acute illness [13].

### Laboratory methods

Upon collection, annual blood samples were immediately transported to the Nicaraguan National Virology Laboratory for processing and storage at -80°C. Paired annual samples (2014-2015 and 2015-2016) demonstrating seroconversion by CHIKV Inhibition ELISA [16] indicated CHIKV infection. ZIKV infection status was confirmed by the 2017 result of the ZIKV NS1 blockade-of-binding assay [12,17] on paired 2017-2018 annual samples.

Suspected chikungunya cases were confirmed by 1) a multiplex pan-DENV and CHIKV real-time RT-PCR (rRT-PCR) on acute blood samples [18], 2) seroconversion using a CHIKV IgM ELISA on paired acute and convalescent samples [16], and/or 3) seroconversion with a *≥*4- fold increase in titers levels on paired acute and convalescent samples as measured by a CHIKV Inhibition ELISA [14,16]. Suspected Zika cases were confirmed by either rRT-PCR in acute blood and/or urine samples or a serological algorithm [4,19] based on acute and convalescent serum samples measured by the ZIKV and DENV Inhibition ELISAs, the IgM-capture ELISAs, and the ZIKV NS1 blockade-of-binding assay. The algorithm was the product of recursive partitioning of classification trees and was cross-validated [19].

### Testing criteria for cases

PDCS participants who were ill and reported to the HCSFV during the study period were tested for an acute, symptomatic CHIKV and ZIKV infection if they exhibited certain clinical profiles. During the two chikungunya epidemics, these clinical profiles were: 1) fever and at least two of the following: headache, retro-orbital pain, myalgia, arthralgia, rash, hemorrhagic manifestations, and leukopenia [the 1997 World Health Organization (WHO) dengue case definition [20]] and 2) undifferentiated fever without evident cause, with or without any other sign, symptoms, or complete blood count finding. The two clinical profiles that constituted the study’s testing criteria reflected the PDCS’s original goal of studying DENV infections. However, they were expansive enough to capture the known manifestations of chikungunya and the official WHO case definition for chikungunya at the time, which was simply fever and arthralgia [21].

When the PDCS expanded to include Zika, one additional clinical profile that triggered arboviral testing was enacted: 3) afebrile rash, with or without any other sign, symptoms, or complete blood count finding. This change to the testing scheme was enacted because early reports [22] and preliminary guidance from the Pan American Health Organization (PAHO) stated that Zika could infrequently present without fever.

### Statistical analyses

We measured the infection incidence (new infections / immunologically naive population at the start of the epidemic) and disease incidence (new cases / immunologically naive population at the start of the epidemic) of each epidemic using intercept-only logistic models with a generalized estimating equation (GEE) framework. (We note that infection incidence is also called seroprevalence, infection rate, and risk of infection; similarly, disease incidence is known as the attack rate and incidence proportion of disease.) Spatial incidence was mapped across the study area with spatial generalized additive models [23] with bivariate spatial splines for the study households’ longitude and latitude, where participants were geolocated. The intracluster correlation coefficient, which measures the intra-household correlation of incidence measures, was estimated using three different estimators based on ANOVA, GEE, and resampling approaches. Using SaTScan, we employed Kulldorf’s spatial scan statistic alongside 999 Monte Carlo simulations per iteration to identify hierarchical and Gini clusters [24,25]. Geostatistical logistic mixed effects models [26] quantified the association of risk factors with infection and disease while accounting for correlated outcomes across space and within households. Their results were compared with standard logistic models (which accounted for no sources of correlation) and logistic mixed effects models (which only accounted for within-household correlation). Infection dynamics were estimated by treating cases as a spatiotemporal Poisson point process arising from the total population and then accounting for the spatial distribution of the proportion of asymptomatic infections, which was assumed to be time-invariant. Analyses were performed in R. Extensive information regarding the statistical analysis is provided in the Supplement.

## RESULTS

### Participant characteristics

Our study assessed the infection and disease status of 3,693 unique PDCS participants across ChikE1, ChikE2, and ZikaE. Approximately 3,000 PDCS participants were analyzed in each epidemic (Table 1), and all participants were uninfected at the start of each epidemic we assessed. These three epidemics occurred in 2014-2016 throughout Managua’s rainy period of June-November (Fig. 1), during which an abundance of mosquitoes is observed in the study area. The distribution of age and sex was similar across ChikE1, ChikE2, and ZikaE (Tables 1 and S1, Fig. S2), with approximately 50% of PDCS participants being female in each epidemic.

**Figure 1.**
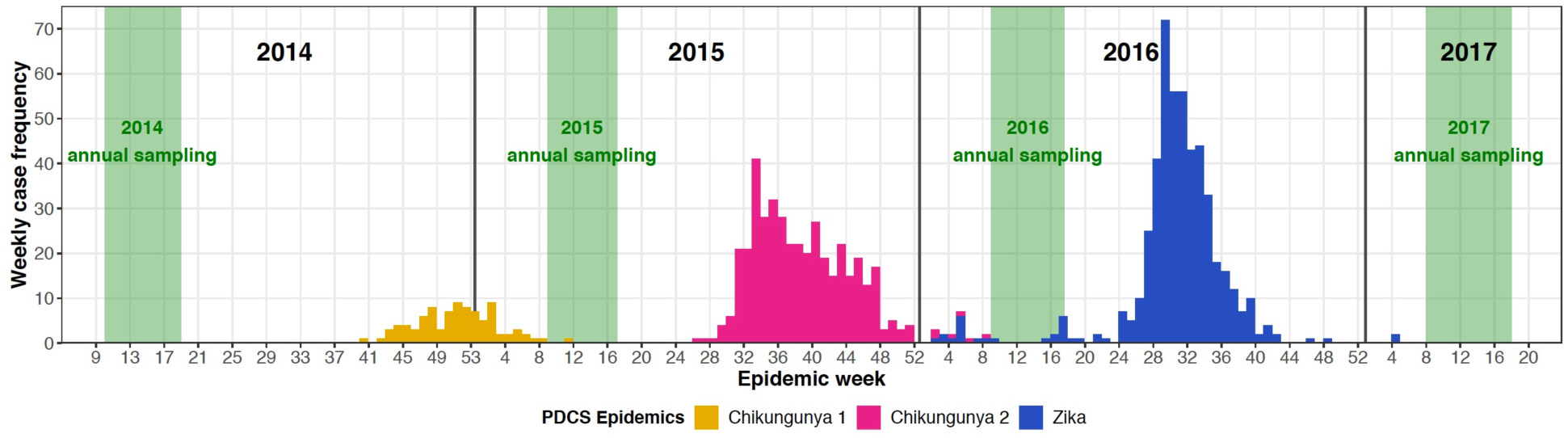
Epidemic curves for three epidemics in the PDCS in Managua, Nicaragua, on a weekly basis. The duration of the annual serosurvey that detects infection status is shown in green. Epidemic curves for the chikungunya and Zika epidemics reflect case counts (numbers of cases) that were confirmed by real-time RT-PCR and a serological algorithm (see Supplement).

**Table 1.** Summary information for three large arboviral epidemics in the PDCS in Managua, Nicaragua*.

|  | **1<sup>st</sup> chikungunya**<br>**epidemic (ChikE1)** | **2<sup>nd</sup> chikungunya**<br>**epidemic (ChikE2)** | **Zika**<br>**epidemic (ZikaE)** |
| --- | --- | --- | --- |
| **Epidemic period** | 9/2014 – 2/2015 | 7/2015 – 2/2016 | 1/2016 – 1/2017 |
| **Number at risk of new**<br>**infection at epidemic's**<br>**start** | 3,124 | 2,864 | 3,017 |
| **Median age (IQR)** | 8.5 (6.0-11.8) | 8.7 (6.0-11.6) | 8.9 (6.2, 11.7) |
| **Female (%)** | 1,571 (50.3%) | 1,441 (50.3%) | 1,516 (50.2%) |
| **Number of new**<br>**infections** | 199 | 710 | 1,416 |
| **Number of**<br>**symptomatic infections**<br>**(cases)** | 90 | 416 | 494 |
| **Infection incidence**<br>**(95% CI)<sup>1</sup>** | 6.4%<br>(5.5%, 7.4%) | 24.8%<br>(23.2%, 26.6%) | 47.1%<br>(45.1%, 49.1%) |
| **Disease incidence**<br>**(95% CI)<sup>1</sup>** | 2.9%<br>(2.3%, 3.6%) | 14.5%<br>(13.2%, 16.0%) | 16.6%<br>(15.2%, 18.1%) |
| **ANOVA-based ICC of**<br>**infection status**<br>**(95% CI)<sup>2</sup>** | 0.22<br>(0.17, 0.28) | 0.21<br>(0.16, 0.27) | 0.22<br>(0.17, 0.27) |
| **ANOVA-based ICC of**<br>**disease status**<br>**(95% CI)<sup>2</sup>** | 0.04<br>(0.00, 0.09) | 0.14<br>(0.09, 0.21) | 0.16<br>(0.11, 0.21) |

\*Abbreviations: ANOVA, analysis of variance; CI, confidence interval; ICC, intracluster
correlation coefficient; IQR, interquartile range; PDCS, Pediatric Dengue Cohort Study
<sup>1</sup> Estimates derive from logistic models using a generalized estimating equation framework.
<sup>2</sup> Table S5 contains additional information.

### Summary measures of infection and disease

We first examined summary statistics of the three epidemics. ChikE1 exhibited the lowest infection incidence at 6.4 cases per 100 population (6.4%) and about half as much disease incidence (2.9%) (Table 1). Although ChikE2 and ZikaE exhibited similar disease incidences (14.5% and 16.6%, respectively, p=0.054), the infection incidence was significantly lower, by about half, for ChikE2 compared to ZikaE (24.8% vs. 47.1%, respectively, p<0.001) (Table 1). Despite occurring in the same population with little year-to-year variation in age structure, socio- economic conditions, etc., the percentage of new infections resulting in disease was significantly higher in ChikE2 than ChikE1 (58.6% vs. 45.2%, p=0.001) (Table 1). ZikaE featured the lowest percentage of symptomatic infections (34.9%) across all three epidemics (p=0.006 compared to ChikE1 and p<0.001 compared to ChikE2).

We then assessed incidence by sex and age. Sex-based differences for incidence measures, even when statistically significant, tended to be small, as when females had an infection incidence 6% higher than males during ZikaE (Fig. S3). In contrast, we observed non-linear age-incidence patterns for all epidemics (Fig. S4), which were higher at every age than and had steeper increases across age than disease incidence (Fig. S5). For example, ZikaE disease incidence was low (10-15%) across early childhood, and it noticeably increased during adolescence to ∼28% among the oldest participants (Fig. S6). However, ZikaE infection incidence was high across all ages, increasing dramatically from ∼28% in infants of 2 years to ∼69% in adolescents 14 years old. As with the overall, epidemic-wide disease incidences for ChikE2 (14.5%) and ZikaE (16.6%), the age-disease incidence trends for these two epidemics were very similar. However, the epidemics’ age-infection incidence trends diverged substantially, with infection incidence being significantly higher at every age during ZikaE compared to ChikE2.

### Spatially mapping infection and disease incidence

Next, we mapped the infection and disease incidences across the study area. For all epidemics, infection incidence varied at small spatial scales (Fig. 2A-F), suggesting that the local environment was an important determinant of infection. During ChikE1, ChikE2, and ZikaE, infection incidence was elevated in western neighborhoods adjacent to a large cemetery that is frequently and heavily infested with *Aedes* mosquitoes during the rainy season. Adjusting Fig. 3A-C for distance to the cemetery appreciably changed the spatial patterns of infection incidence, whereas adjusting for age, sex, and household water availability did not (Figs. S6- S11).

**Figure 2.**
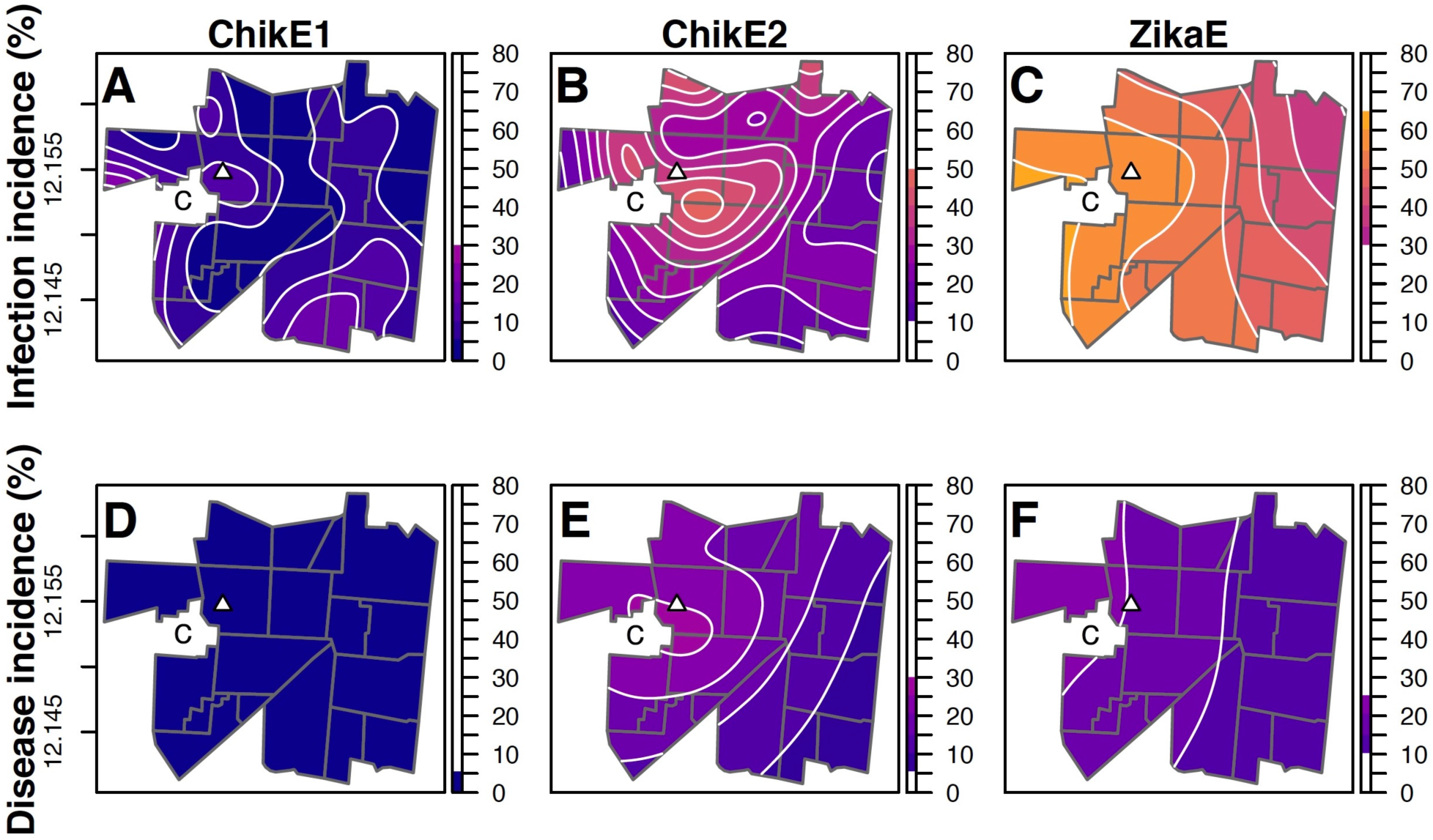
Maps of the infection incidence and disease incidence. The infection incidence (A- C) and disease incidence (D-F) across three epidemics are shown in one color palette to facilitate visual comparisons, with warmer colors indicating higher values. Contour lines show changes in incidence discretized into bins for every 5 percentage points and correspond to the scale bar of color-coded values. Maps were generated from generalized additive mixed models. The white triangle indicates the location of the study health center. Neighborhoods are outlined in gray. The location of the cemetery is marked by the letter “C.”

**Figure 3.**
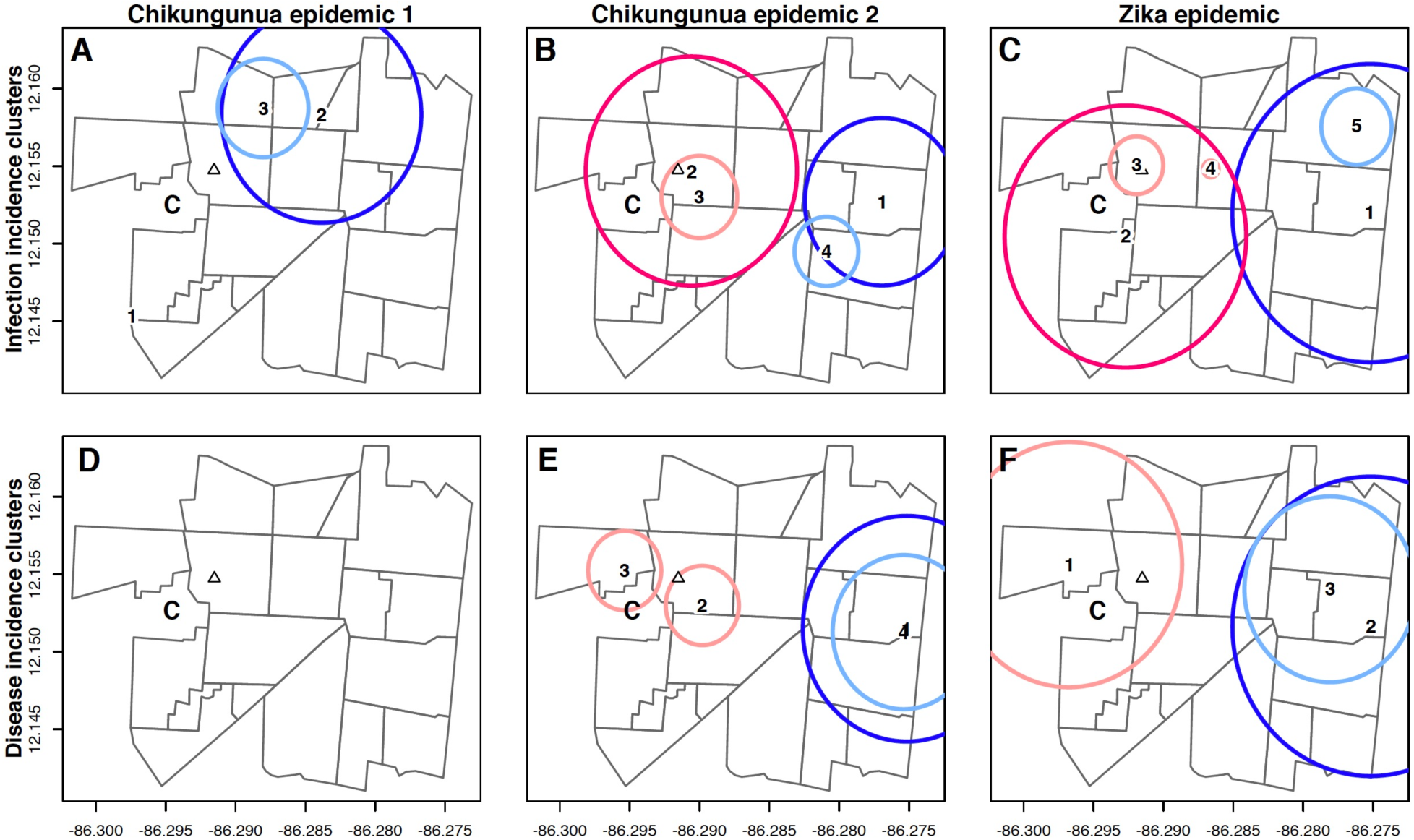
Cluster detection analyses of the infection and disease incidence. Clusters of infection incidence (A-C) and disease incidence (D-E) across three epidemics are shown. Panels depict the results of Kulldorf’s spatial scan statistic conducted in SaTScan. Hierarchical clusters are shown in dark colors; Gini clusters are shown in light colors. Hierarchical clusters identify the most statistically likely clusters; Gini clusters maximize incidence measures. Hotspots are shown in pink; coldspots are shown in blue. Cluster centers are numerically labeled. For panel E, the clusters 1 and 4 have nearly identical centers, so their label numbers are superimposed on each other. The white triangle indicates the location of the study health center. Neighborhoods are outlined in gray. The location of the cemetery is marked by the letter “C.” Table S2 contains additional information for this analysis.

Across all epidemics, disease incidence also varied at small spatial scales (Fig. 2D-F), but both its variation and magnitude were noticeably smaller compared to the spatial distribution of infection incidence. When averaging over the predicted incidences across space, the largest differences between the disease and infection incidences was observed for ZikaE, which featured a disease incidence 34.9 percentage points smaller than infection incidence. After adjustment (Figs. S12-17), spatial patterns of disease incidence remained distinct from spatial patterns of infection incidence. Together, these observations demonstrate that for ChikE1, ChikE2, and ZikaE, infection incidence had a different spatial pattern, magnitude, and variation than disease incidence.

### Cluster detection

We then identified hierarchical and Gini clusters of infection and disease incidence (Fig. 3, Table S2). These clusters can identify areas of both excess and deficient incidence relative to the background, study area-wide incidence. Each epidemic had ≥1 significant cluster. Clusters of elevated infection incidence for the larger epidemics, ChikE2 and ZikaE, encompassed the westward cemetery and study neighborhoods adjacent to it. These clusters of elevated infection incidence denoted areas with excess and newly infected persons during each epidemic.

Conversely, large clusters of diminished infection incidence in ChikE1, ChikE2, and ZikaE highlighted areas in the eastern part of the study area with excess uninfected persons who remained susceptible to future infection. Standard disease incidence clusters, which identify areas of elevated or diminished case counts among the total population, were observed for ChikE2 and ZikaE; however, they were not observed for ChikE1 despite the detection of a large infection incidence cluster in the northern half of the study area (Fig. 3).

### Geostatistical modeling

Using three independent methods, we found surprisingly low levels of both correlated infection status (intracorrelation coefficients ≈ 0.2) and disease status (intracorrelation coefficients ≈ 0.1) within homes (Tables 1, S3-S5; Figs. S18) for all epidemics. Using geostatistical models, we accounted for potential correlation of infection and disease status within homes and across space. With these models, the strongest signal we observed indicated that distance to the cemetery was significantly associated with infection incidence during ZikaE, such that the odds of ZIKV infection among participants living 1 km from the cemetery were 0.63 (95% CI: 0.55, 0.73) times that of participants living next to the cemetery, conditional on age, sex, and indoor water availability. A similar 1-km odds ratio (OR 0.65; 95% CI: 0.55, 0.77) was observed during ChikE2 (Table S3). Age was significantly associated with both infection and disease incidence for all three epidemics (Tables S3-4). While female sex was not associated with disease incidence during the chikungunya epidemics, it was positively associated with disease incidence during ZikaE (OR 1.29; 95% CI: 1.05, 1.58). Conversely, prior DENV infection was negatively associated with disease incidence during ZikaE (OR 0.77, 95% CI: 0.61, 0.99).

A comparison of models that did and did not account for spatial correlation of infection and disease status revealed nearly identical point and interval estimates (Tables S3-4), suggesting that spatial correlation in our study setting operated over short distances. This was confirmed by graphing the autocorrelation function by epidemic (Figure S19). For the Zika epidemic, it showed that the ZIKV infection status of participants living >100 meters apart had an average correlation <0.05. Similarly, the CHIKV infection status during ChikE1 and ChikE2 of participants living >200 meters apart had an average correlation of <0.2.

### Spatiotemporal dynamics

Spatiotemporal analyses depict epidemic progression across time and space. We estimated the spatiotemporal dynamics of infection across six months for each epidemic (Fig. 4A-C). The pattern of infections indicated that ChikE2 and ZikaE began along the western edge of the study center next to the large cemetery where mosquitoes frequently breed during the rainy season.

**Figure 4.**
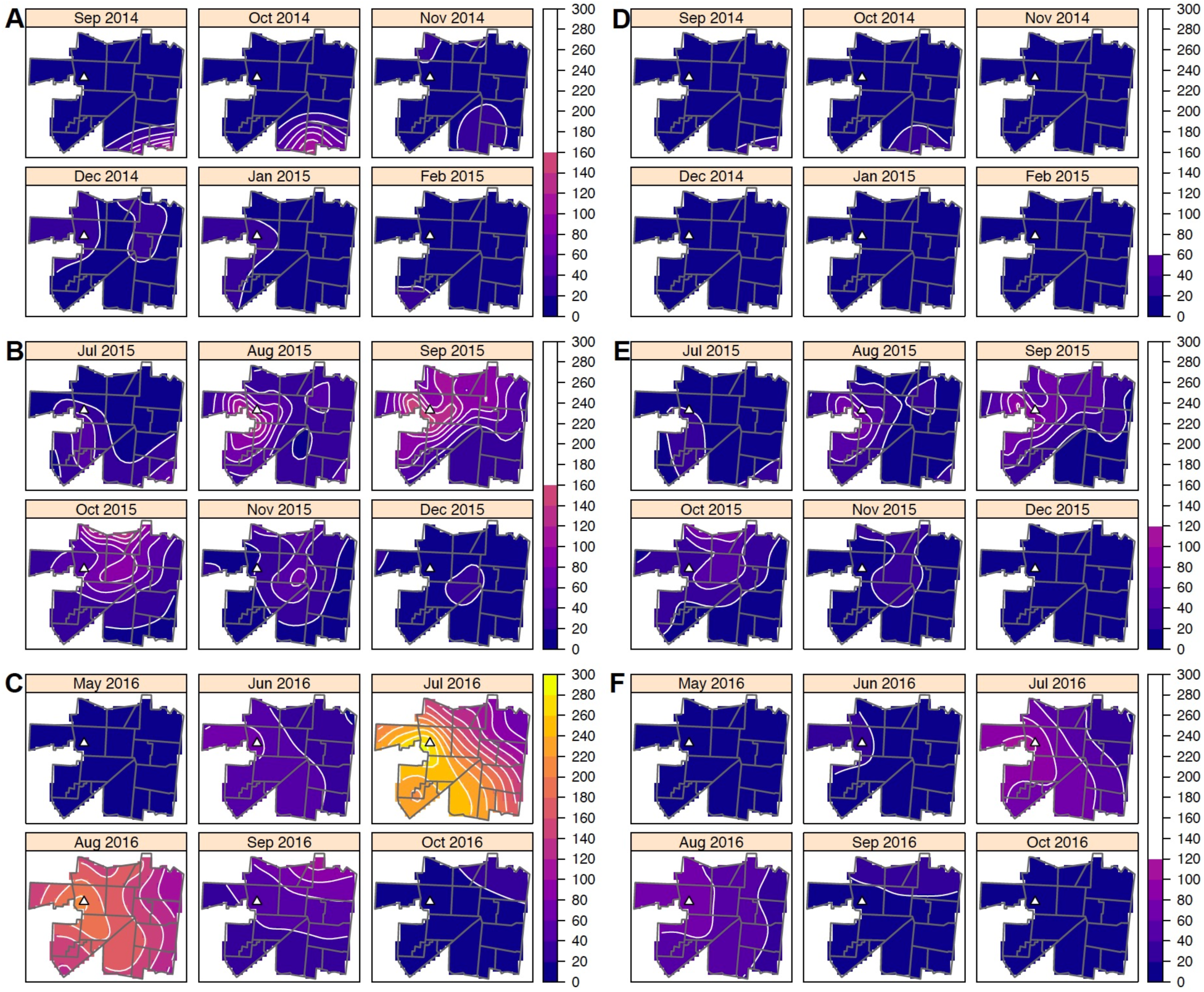
Spatiotemporal dynamics across three epidemics in our study area. Model predictions of the infection incidence (A-C, first column) and disease incidence (D-F, second column) are reported per-month and per-1,000 population in the PDCS. Due to space constraints, data for months with only a few cases are not shown. Maps for ChikE1, ChikE2, and ZikaE are shown in the same color palette and use the same color binning to ease cross-epidemic comparisons. Contour lines show changes in incidence discretized into bins for every 20 cases per 1,000 population and correspond to the scale bar of color-coded values. The white triangle indicates the location of the study health center. Neighborhoods are outlined in gray. Rows in the figure correspond to ChikE1 (A, D), ChikE2 (B, E), and ZikaE (C, F) in Managua, Nicaragua, from top to bottom.

Conversely, the smaller ChikE1 began in the southeastern corner of the study area where no known locus of mosquito activity has been identified. Nevertheless, for all three epidemics, elevated infection incidence around cemetery-adjacent neighborhoods was evident for at least two months, with particularly high intensity during ZikaE. High infection risk around the cemetery early in ChikE2 and ZikaE was followed by a shift toward northern, lake-adjacent neighborhoods; this pattern was not observed during ChikE1. In contrast to the spatiotemporal observations of infection, the spatiotemporal dynamics of disease incidence were muted across epidemics (Fig. 4D-F). The spatiotemporal pattern of infection incidence for ZikaE (Fig. 4C) differed most from its pattern of disease incidence (Fig. 4F).

## DISCUSSION

Here we describe the spatial and spatiotemporal dynamics of two large chikungunya epidemics and one explosive Zika epidemic in a pediatric cohort in Managua, Nicaragua. We demonstrate that areas of high infection incidence, whether identified directly via smoothed estimates or coarsely via cluster analyses, were not necessarily areas of high disease incidence, and vice versa. We consistently found large clusters of infections, but small if any clusters of disease, suggesting that infection but not disease is the spatially clustered phenomenon for CHIKV and ZIKV in our study site. Among our participants, arboviral infections were correlated across short distances, very weakly clustered within households, and were not associated with indoor water availability, suggesting that non-household transmission drove the three epidemics. Multiple analyses implicated the local cemetery as a major source of infections in all epidemics; however, the cemetery was likely a major source of epidemic initiation for ChikE2 and ZikaE, as ChikE1 appears to have begun in the southeastern part of the study area. By understanding the spatial distribution of infections, not just cases, it becomes possible to improve targeting of future interventions.

Surprisingly, and in contrast to a general [5,6,9] (though not universally observed [7,8]) assumption in arbovirology, we found little evidence across three major epidemics that infections clustered within households. These results are consistent with our prior study of full-length sequencing of ZIKV genomes in the PDCS [8], which found that most study households had Zika cases whose most recently sampled viral ancestral strains derived from a different household. Indeed, our prior research showed that the majority of first-generation transmission events during ZikaE were among individuals who lived over one kilometer apart. The weak household correlation of infection status among children who lived together during all three epidemics is also consistent with a prior measure of the intracluster correlation for infection status (∼0.1) among pediatric and adult household members during ChikE2 [11]. The available evidence suggests that non-household transmission played an important role in the chikungunya and Zika epidemics we assessed. On this basis, we hypothesize that non-household transmission may also be the primary driver of dengue virus transmission in our cohort. Future studies will be needed to interrogate this hypothesis and compare dengue transmission to our chikungunya and Zika results. The large cemetery abutting our tropical study area has ∼22,250 single-body graves, multi-body crypts, and mausoleums. The typical grave is adorned with two cement vases and a deep recess for a coffin-length garden. These embellishments fill with rainwater and create highly conducive environments for *Aedes* mosquito breeding [27], likely explaining why participants living near the cemetery exhibited high infection incidence during all three epidemics. Study entomologists have previously found very high numbers of larvae, pupae, and adult *Aedes aegypti* mosquitoes in this cemetery (H. Suazo, personal communication). Areas of elevated infection risk were also observed in the Julio Buitrago neighborhood; site visits by the study team’s entomologists in 2018 revealed several shops that sell and repair tires, which capture rainwater in their interior and promote mosquito breeding. Given the lack of evidence for household transmission, assessing the built environment and key non-household sites for mosquito breeding sites may help design interventions to curtail future epidemics.

Distinct from many spatial studies of arboviruses [28–33], our analyses separately considered infection and disease as we had access to both variables in a large, prospective cohort. Without data to the contrary, it is reasonable to assume that the spatial pattern of disease incidence, which is measurable with case data, maps reasonably well to the spatial pattern of infection, which can only be determined by capturing both symptomatic and asymptomatic infections. However, for all three epidemics we assessed, we found that the spatial patterns of infections were distinct from those of disease. This implies that among the infected population in our cohort, the proportion of individuals exhibiting disease may vary considerably over space. A companion study explores this possibility in greater depth than is possible here.

Most spatial analyses of chikungunya and Zika epidemics have used areal data [34–40] (information grouped at a neighborhood, city, or district level), as more granular point data is difficult, but possible [41,42], to obtain at large spatial scales. Studies using areal data may be impacted by the ecological fallacy (inferring that associations at the group level apply to the individual level) [43] and its spatial counterpart bias, the modifiable areal unit problem (the aggregation of data into arbitrary spatial units) [44]. Our study design and the available PDCS data limited these biases from impacting our findings.

Separate from the main results of our spatial analyses, we note that our previously published estimate of the infection incidence during ZikaE was 36.1% in the PDCS [14]. However, we now revise this estimate here to 47.1%. The increase is primarily due to an improved, second-generation NS1 blockade-of-binding assay used to detect ZIKV infections and the use of a multi-assay algorithm that captured rRT-PCR-negative, serology-positive Zika cases [4,19].

Our study was additionally strengthened by the prospective cohort study design, the large number of participants, reliable measurements of infection status, a case ascertainment strategy capable of capturing very mild illnesses [3,4], and the lack of pre-existing immunity to CHIKV and ZIKV in our study population. However, the study has several limitations. The pediatric nature of the PDCS precluded spatially analyzing adults in the catchment area of the study health center during ChikE1, ChikE2, and ZikaE. However, previous analyses in the area found that the spatial pattern of infection incidence during ZikaE was similar for children and adults, though the magnitude of incidence was higher among adults [12]. Thus, analyses of the adult population during ChikE1 and ChikE2 may have revealed similar spatial trends as those in PDCS participants. In addition, the geographic extent of our study is small (5 km^2^), which restricted the number of infections and cases we observed. However, capturing all infections, especially asymptomatic ones, is only cost-feasible in constrained geographical areas.

In summary, our results demonstrate that three major epidemics of chikungunya and Zika within the PDCS exhibited spatial and spatiotemporal similarities as well as epidemic-specific differences. Across all three epidemics, we found that areas of high infection incidence did not necessarily have high disease incidence. Consistent evidence emerged that Managua’s general cemetery is a key feature of chikungunya and Zika epidemics in the PDCS. Our data support public interventions in non-household transmission sites as a critical approach to decrease arboviral epidemics. As *Aedes* mosquitoes are predicted to expand their ecological range in the coming decades [45], aligning local mosquito control strategies to the spatial contours of infections will strengthen epidemic management.

## ACKNOWLEDGMENTS

We are extremely appreciative of our dedicated study team at the Centro de Salud Sócrates Flores Vivas and the Laboratorio Nacional de Virología at the Centro Nacional de Diagnóstico y Referencia, Nicaraguan Ministry of Health; and the Sustainable Sciences Institute in Nicaragua. We thank Douglas Elizondo for the management of cohort databases. We are grateful to Art Reingold and Tulika Singh for their thoughtful reviews of the manuscript, and we thank Burke Bundy and Suzanne Default at the University of California, Berkeley, for enabling us to use the computer cluster of the Division of Epidemiology and Biostatistics for our geostatistical modeling. We thank François Rousset for expert consultation regarding spatial generalized linear mixed models and their implementation in the spaMM R package. Most importantly, we thank the PDCS participants and their families for engaging with us in the endeavor of science.

## AUTHOR CONTRIBUTIONS

FBC and EH conceived the study. AB, KG, and EH developed the study design. SO, NS, MP, HSL, AB, and KG implemented the study design and collected field data. AB developed and supervised the laboratory assays. SS, DC, and TM performed laboratory testing. BLM, JCM, DE, and SA cleaned and verified the data. FBC analyzed the data and generated the figures. FBC, HS, and EH drafted and revised the manuscript, and all authors reviewed the manuscript. FBC, AB, and EH acquired funding.

## GENERATIVE AI USE STATEMENT

The authors report generative AI was not used in their research or preparation of this manuscript.

## DATA AVAILABILITY STATEMENT

Individual-level data can be shared with investigators after approval by the University of California, Berkeley Institutional Review Board (IRB). Data will be available upon request (as required by IRB-approved protocols for the PDCS) to the University of California, Berkeley Center for the Protection of Human Subjects and Eva Harris to arrange for data access. Collaborating research groups and institutions will be sent coded data with all personal identifiers unlinked including the data dictionaries and accompanying R code.

