## Supplemental materials for "Spatial analyses of infection and disease during chikungunya and Zika epidemics in Managua, Nicaragua"

### Supplementary materials and methods

#### Statistical and data visualization considerations

*GPS coordinates*

Nicaragua does not use a house numbering system, whereby every home in a given city is assigned a unique number along a street to define the household address. Instead, addresses are officially recorded with reference to common landmarks (*e.g.,* the large tree in the Mantica neighborhood) and geographical aspects (*e.g.,* three blocks toward the lake and one block up [East]) of a city. The address of each Pediatric Dengue Cohort Study (PDCS) household is recorded upon participant recruitment into the cohort study and confirmed at subsequent annual samples. When a participant moves to a new house, the new address and the date of the move is logged in the cohort study's computer systems. Global Positioning System (GPS) points are taken upon site visits to the approximate 2,000 PDCS households for annual field visits, follow-up questionnaires, and medical checks at the household, as needed. The annual field visits serve to encourage continued participation in the PDCS, receive feedback from participants, collect annual samples for participants who did not visit the study health center on time, and survey participants regarding their use of both study and non-study health centers for medical attention.[1] Household address and GPS information for all members of the PDCS for all years we considered were systematically examined, validated, and geolocated using the EPSG:4326 coordinate reference system by a group of study authors who live in Managua and are extensively familiar with the study area. The same reference system was used for spatial analyses.

Individuals who lived in the same household were initially assigned the same GPS points. However, some statistical methods we used are not designed to work with different individuals who have identical GPS points; doing so would cause some of the algorithms we used to return an error. We therefore slightly jittered the points of all study participants. The average displacement (distance from original GPS point to jittered GPS point) was 0.09 meters (3.5 inches), with the maximum displacement being 0.16 meters (6.5 inches). The jittering was performed using the jitter function in R, which added a uniform amount of noise to the original GPS points.

*Household correlation*

During ChikE1, ChikE2, and ZikaE, approximately 35-40% of PDCS households had a single participant; thus, most households had more than one participant. Correlation of infection status at the household level, due to shared environments, genetics, and behaviors, was therefore possible. This could lead to a violation of the independence assumption embedded in standard regression approaches and other methods we used. The possibility of household-based correlation necessitated the use of techniques that could account for or measure this across many of the analyses we performed. As detailed further below, this was accomplished through calculation of the intra-cluster correlation coefficient, mixed-effects geospatial models, generalized estimating equations, and generalized additive models with random effect spline terms. All analyses accounted for household-based correlation, unless otherwise noted.

*Generalized linear models*

Generalized linear models (GLM) generalize linear regression (the general linear model) by allowing the linear predictor (systematic component) to be related to possibly non-continuous outcomes through a variety of link functions. Instead of directly modeling the outcome variable as in linear regression, a GLM models a function of the mean of the outcome variable. We used logistic regression (a GLM with a logistic link function and an assumed binomial distribution for the outcome variable [random component]) to estimate odds ratios and corresponding 95% confidence intervals (CIs) for factors plausibly related to infection and disease status.

The GLM estimation framework returns unbiased measures of association under a critical assumption of independence. In our study setting, this assumption may be violated two different ways: by correlated data among PDCS participants living in the same household and/or by correlated data among participants living close to each other in space. Both possible sources of bias necessitated the use of more complex models.

*Generalized estimating equation models*

Models using generalized estimating equations (GEE)[2] extend GLMs to account for more complex data structures, including correlated data. Epidemic-specific, intercept-only logistic GEE models were used to calculate the incidence of infection and the incidence of disease. GEE models were run using the geepack R package[3] with an exchangeable correlation structure, Huber-White standard error estimators,[4] and the scale parameter for a generalized binomial distribution fixed to 1. Household-based clustering was dealt with by setting the household ID variable as the clustering variable, such that infection and disease status across households are assumed to be independent. Model results were backwards-transformed from the logit scale to the probability scale. Estimation of infection incidence accounting for household-based clustering (Figure S18) was performed with a GEE model instead of a mixed-effects model because estimates from logistic GEE models, unlike logistic mixed-effects models, are interpretable as population-level averages across the clustering variable.[5,6]

The risk difference is the difference between two risks (proportions). For each epidemic we considered, we calculated the risk difference using the incidence of infection among male and female participants. The same was done with the incidence of disease. To account for household-based clustering in estimating risk differences, we used modified Poisson regression within a GEE framework (Poisson distribution, the identity link function, the household as the clustering variable, and robust standard errors[7,8]).

*Generalized additive models*
Generalized additive models (GAMs)[9,10] are semi-parametric extensions of GLMs that fit smooth functions to the data to account for possibly non-linear trends. GAMs are particularly well-suited to model continuous variables, such as exact age or GPS coordinates, because they can capture complex, non-linear relationships that would be missed by standard GLMs. All GAMs were estimated with the mgcv R package[10] GAMs used restricted maximum likelihood (REML), as recommended,[11,12] to estimate the optimal smoothing parameters,[12,13] and used the outer, Newton numerical optimization method for smoothing parameter estimation. Thin plate splines were used. Household-based clustering was dealt within the GAM estimation framework by including a random-effect penalized smoothing basis on the household ID variable. Model predictions from the GAMs to the probability scale excluded the random effect terms, so that they were zeroed out in the prediction process.

Logistic GAMs were used to assess the overall trend of exact age with the incidence of infection and disease. Spatial logistic GAMs were used to estimate the incidence of infection and disease for the spatial extent of the study area. Spatial logistic GAMs (for both contour and perspective plots) used a bivariate spatial smooth (two-dimensional spline) that took the jittered longitude and latitude coordinates for a given individual as the input. The contour and perspective plots display the predicted probability of the outcome after holding the relevant covariates constant at their median values across all epidemics. The use of a single set of median values for contour and perspective plots allow for cross-epidemic and cross-plot comparisons for PDCS data. For ease of comparisons, all perspective plots were visualized at an azimuthal angle of 47.5º and a colatitude angle of 30º.

*Intracluster correlation coefficient*
The intracluster correlation coefficient (ICC) was used to measure the intrahousehold correlation of infection and disease status as it measures the similarity of outcomes within clusters. The ICC is calculated as the ratio of the between-cluster variance to the sum of the within- and between-cluster variance. The value of the ICC ranges between 0 and 1. An ICC of 0 indicates that outcomes within the cluster (participant households) are uncorrelated with each other. An ICC of 1 indicates that outcomes within a cluster have the same outcomes as each other. If households served as the site of viral transmission, we would expect the ICC to be closer to 1 (reflecting an abundance of either all-infected households or all-uninfected households) than 0. To not bias the ICC by the inclusion of households that only have 1 participant (and hence a mathematically degenerate within-household variation), only households with $\geq$2 participants were used to estimate the ICC. However, when including all households, results were very similar.

There is a vast literature on different ICC estimators' properties,[14–16] and we aimed to demonstrate that our results were robust to the specification of the ICC estimator. First, we estimated the ICC using the traditional one-way ANOVA approach using Searle's exact confidence limit equations[17] and the ICCest() function of the ICC R package[18]. Second, we used a multivariable logistic GEE model with an exchangeable correlation structure to measure the ICC. The variance of the correlation coefficients were extracted from geese() function output, specifically the valpha term. GEE models for CHIKV and ZIKV infection adjusted for exact age, sex, water availability, and distance to the cemetery. GEE models for chikungunya and Zika occurrence conditioned on exact age and sex for all comparisons. The GEE models for Zika occurrence further conditioned on prior DENV infection status for participants examined during ZikaE. Third, the ICC was estimated using the modern approach of Chakraborty and Sen,[19] which is based on resampling and U-statistics, using the iccbin() function of the ICCbin R package[20] and a resampling-based approach for the confidence intervals.

*Household infection risk vs household size*
If households functioned as major sites of CHIKV and ZIKV transmission, then household-level infection incidence could be larger for households of larger sizes, as larger households would have more opportunities for mosquito breeding grounds and a larger concentration of individuals that mosquitoes could feed on.[21,22] To examine this possibility, we plotted household size vs. household infection risk (Figure S18). We plotted the population-averaged, intercept-only GEE estimate of infection risk, which averages the estimated infection incidence across households of different sizes. We then overlaid the result of a LOESS (locally estimated scatterplot smoothing) regression[23,24] to show the underlying trend in the data. The span of the LOESS algorithm was set to the default value of 0.75.

*Kulldorff's spatial scan test*
Kuldorff's spatial scan test[25] was used to detect spatial clusters based on a significant excess or deficit of outcomes (whether infections or cases) within a moving window. This window visits all spatial locations and varies in size to detect small and large clusters. A purely spatial analysis was conducted with a Bernoulli probability model within SaTScan.[26] Clusters of high and low rates were scanned for with a circular spatial window. The maximum allowable size of a cluster was set at 50% of the population at risk (the default) for the outcome of interest, and 999 Monte Carlo simulations were run to generate p-values. SaTScan adjusts p-values for the millions of multiple comparisons it conducts. By default, SaTScan maximizes the likelihood to identify hierarchical clusters, which are therefore the most statistically likely clusters. Hierarchical clusters are the most common type of clusters that are reported in spatial health research. They can be any size, but they tend to be large and may miss smaller, true clusters. As a result, we also used the SaTScan option to identify clusters by considering the Gini index.[27] These Gini clusters maximize outcome rates when comparing cluster and non-cluster areas. Due to the way Gini clusters are calculated, they tend to be smaller than hierarchical clusters, but they have higher rates of the outcome of interest.

Statistically significant, non-geographically overlapping hierarchical clusters as well as significant Gini clusters were visualized, as is the norm in spatial health research. To identify clusters of high and low infection incidence, we compared infected (cases) vs. uninfected (controls) participants. (Here we use case-control nomenclature that is standard in the cluster detection literature. This nomenclature assigns a different meaning to *case* that does not imply a symptomatic infection, as we use *case* throughout most of the text. See Table S2 for further details.) To identify clusters of high and low disease incidence, we compared symptomatic infections (cases) vs. uninfected persons and those with subclinical infections (controls). This method does not account for household-based clustering.

*Moran's I test*
Global Moran's I[28] test was used as an intermediary step to assess whether outcomes were spatially correlated (spatially autocorrelated). For Moran's I test, deviance residuals from non-spatial, logistic mixed-effects models were used as the numerical vector to assess whether outcomes remained autocorrelated after adjusting for covariates. The weights for Moran's I test corresponded to inverse distances between participants' locations. Holistically considering the output from Moran's I test, we proceeded with using spatial regression models.

*Statistical models for estimating measures of association*
A series of regression models were constructed to estimate odds ratios of risk factors, chosen *a priori*, related to each outcome of interest. This was done to evaluate the impact on point and interval estimates after accounting for household-based clustering and spatial correlation. First, standard logistic GLMs were built, which account for neither clustering nor for autocorrelation.

Second, logistic generalized linear mixed models (GLMMs)[29,30] that account for household-based clustering were built by including the household ID as a random intercept. GLMMs are flexible extensions of GLMs that can account for a variety of clustered and longitudinal data structures, similar to GEE models. However, GLMMs account for clustering through the use of random effects. GLMMs were estimated using the lme4 R package.[31] GLMMs were fit by maximum likelihood approaches and used 25 Gauss-Hermite quadrature iterations for parameter estimation, as adaptive Gauss-Hermite quadrature approximation is appropriate for one random effect and is a more accurate approach than the default Laplace approximation. Unless otherwise noted, we used the default bobyqa optimizer for the first phase of optimization (random effects parameters only) and the Nelder-Mead optimizer for the second phase of optimization (random effects and fixed effects parameters). To avoid model convergence issues, we ran some models using the bobyqa optimizer for both optimization steps; when this was done, the maximum number of function evaluations to try was set to 100,000. GLMMs were used to estimate measures of association instead of GEE models because spatial versions of GLMMs exist. In addition, GLMs, non-spatial GLMMs, and spatial GLMMs share an overarching likelihood-based statistical framework that GEE models do not.

Third, spatial GLMMs were implemented with the spaMM R package.[32] Spatial GLMMs are geostatistical extensions of GLMMs that accommodate spatial correlation by modeling autocorrelation directly. Logistic spatial GLMMs were constructed with a random intercept on the household ID variable. The spatial structure of our observations was modeled with a Matérn covariance function on the jittered longitude and latitude variables. Laplace maximum likelihood approximation was used to estimate the correlation parameters and $\lambda$, the variance term of the random effects. Fixed effects were estimated by $h$-likelihood approximation using the penalized quasilikelihood (PQL),[30] specifically the variant that does not use the leverage corrections of REML (PQL/L); by circumventing these corrections, PQL/L uses a marginal likelihood approximation for the estimation of the dispersion parameters. For fixed effect estimation, the PQL/L method was chosen over the default Laplace approximation due to concerns that the former (including its second-order variant[33]) could lead to separation, introducing bias into the estimates.[32] For nugget estimation, the initial value of the parameter was set to 0.5. We constructed spatial GLMMs both with and without accounting for household clustering. We used spaMM's MaternCorr() function to visualize the estimated spatial correlation of two points across distance. For this analysis, infection status was the primary outcome since CHIKV and ZIKV are spread by *Aedes* mosquitoes, which have a limited flight range that can be compared to the extent of spatial correlation for infection status. (The literature contains heterogenous estimates for the mean linear distance that *Aedes aegypti* mosquitoes can travel in their natural environment. However, as recent literature review estimated this distance to be 106 meters.[34]

*Covariate selection*

The covariate set for the regression models were determined by our knowledge of CHIKV and ZIKV, prior literature regarding infection and disease, and our knowledge of the geographical conditions of the study area. The covariate set for CHIKV and ZIKV infection was comprised of exact age, sex, estimated number of hours per day without tap water in the household, and distance (in 100m) from the slightly jittered GPS points to the closest boundary of the polygon representing a local cemetery. The covariate set for disease included age and sex for chikungunya and Zika model; the covariate set for Zika models additionally included previous infection status with DENV.

Exact (fractional) age was based on participants' birthdays and the date that participants last provided an annual sample within a given epidemic period. Age and sex were included in the covariate set as previous work has shown differences in CHIKV and ZIKV infection in children.[35,36] Anecdotes from Managua-based study authors as well as prior studies[37] suggested that the cemetery abutting our study site may serve as a breeding ground for mosquitoes and hence a source of CHIKV and ZIKV infection. A site visit by the lead author in August 2017 confirmed that many of the vases, gardens, water containers, artificial containers, crypts, and other structural aspects of the cemetery served as mosquito breeding sites. We included distance to the cemetery in 100-meter units in the infection models as we hypothesized that mosquito-driven infection incidence would be a function of Euclidean distance for the PDCS epidemics.

Entomological inspections of the shoreline of Lago Xolotlán (Lake Managua) by the Department of Entomology of the Nicaraguan Ministry of Health have revealed breeding sites of *Anopheles* (*albimanus* and *pseudo punctipennis*) and *Culex quinquefasciatus* mosquitoes, but no *Aedes* breeding sites. *Anopheles* mosquitoes commonly found in North America are not known to transmit or be infected by ZIKV;[38] similarly, *Anopheles* mosquitoes are not known to be infected by CHIKV.[39] *Culex* mosquitoes are not thought to be competent vectors of ZIKV[38,40] or CHIKV. As a result, we did not include distance to the lake in the infection models for CHIKV and ZIKV.

We hypothesized that water availability might be related to CHIKV and ZIKV infection incidence as households with low water availability in Managua often store intermittent piped water and rainwater in barrels. These barrels can become mosquito breeding sites if the container walls are not scrubbed, if the water is not frequently replaced, or if larvicide is not used to control larvae growth.

Research has identified existing anti-DENV antibodies from a prior DENV infection as lowering the risk of a subsequent ZIKV infection becoming symptomatic.[41–43] This cross-protection is likely due to the similar immune responses induced to different flaviviruses, particularly cross-reactive antibody responses and/or cross-protective T-cell mediated responses.[41] Because there is no known cross-reactivity between CHIKV (an alphavirus), and DENV (a flavivirus), we only included prior DENV infection in the covariate set for the disease models of Zika.

To our knowledge, there is no existing work showing that distance to cemeteries or other mosquito hotspots influences whether a CHIKV or ZIKV infection becomes symptomatic. However, we added distance to the cemetery in contour plots of the incidence of chikungunya and Zika to examine the hypothesis that the spatial variation in disease incidence is related to elevated viremia from repeated mosquito bites, which are likely to occur close to the cemetery. Direct adjustment for viremia was not possible because viremia levels during acute infections are only measured for cases.

Previous work has shown that elevation is related to arboviral infection incidence.[44] However, as the elevation gradient across our study area is small (42-106m above sea level), we did not believe that household elevation would substantially impact the distribution of the mosquito population, and hence infection incidence, in our study area.

Historically, the household questionnaire that is administered during the annual sampling has been used to construct a validated, principal-components-based proxy variable for socioeconomic status (SES).[45] For the construction of the SES proxy, the following variables are used: whether the household floor is made of earth, whether the household walls are made of concrete, the number of refrigerators/freezers in the household, the number of fans in the household, the number of televisions in the household, and whether someone in the household owns a car or motorcycle. However, due to improving economic conditions over the years, household wealth as measured by this method has become relatively homogenous, including during the years of the present study, which explains earlier null results for the association of ZIKV infection incidence and SES.[35] Thus, we did not include a proxy variable for SES in any model.

Based on surveys collected from PDCS families regarding their use of medical services from the study health center,[1] as well as the results of the clustering analyses for disease outcomes, we saw no evidence that treatment-seeking behavior spatially varied among participants across the three epidemics. Thus, we did not adjust for this factor.

*Spatiotemporal GAMs*

Temporal data were only available for the subset of participants in each outbreak who were cases, as they reported to the health center when they were sick with chikungunya or Zika. Case data were treated as a Poisson point process to investigate the overall spatiotemporal dynamics of cases and infections among the overall population.[46] In particular, cases were treated as a realization from an intensity surface, and the overall study population was used as the offset since its size remained constant across the various months of the epidemic periods.

Cases' reported date of illness onset was used as the temporal variable. For each epidemic, a data frame of case data (*i.e.*, date of illness onset, jittered longitude, jittered latitude, and coordinate reference system) was turned into an sf object using the sf R package,[47] which was then used as the points argument in the space_time_ppmify() function of the disarmr R package.[48] The spatial distribution of the overall study population was aggregated on a 33x33 raster, and this raster was used as the exposure argument in the space_time_ppmify() function. The periods argument, defining the time slices of the analyses, corresponded to a vector of the first date of each month for the duration of a given epidemic period. The approximate number of integration points was set to 10,000 and the output returned a rasterStack for prediction purposes. Poisson regression was applied to the resulting data frame, using the log of the exposure as the offset and the regression weight supplied by space_time_ppmify(). Poisson regression was performed using the bam() function of the mgcv R package,[10] with a tensor product on the bivariate (two-dimensional) spatial smooth given by participants' longitude and latitude and a univariate (one-dimensional) smooth on time period as measured by the month of participants' illness initiation. Thin plate and cubic regression splines were used, with 50 and 5 basis functions, for these two- and one-dimensional smooths, respectively. Predictions for case counts across the raster for the overall population were made for each month and visualized as level plots. The predicted number of cases were visualized per 1,000 persons in the overall study population.

To estimate the spatiotemporal dynamics of infection risk, we used the space_time_ppmify() function to make a purely spatial Poisson model for the number of cases among the infected population. The resulting raster was stacked to create a RasterStack object of the same number of rasters as the months of spatiotemporal predictions for the corresponding epidemic. To back-calculate the spatiotemporal dynamics of infection incidence, we dividing the original spatiotemporal predictions of case occurrence among the overall study population by the RasterStack among the infected population. The predicted number of infections were visualized per 1,000 persons in the study population. This approach assumes that the probability of disease among the infection population varies across space but is constant in time. While this assumption was untestable with the available data, we judged it probable that factors influencing disease occurrence were temporally stable over the short epidemic periods we considered.

*Data visualization*

For all spatiotemporal maps, neighborhood boundaries and contour lines were superimposed on model predictions for ease of interpretation. When possible, data were visualized with the ggplot2 R package[49,50] with uncertainty intervals (pointwise 95% CIs or confidence bands, as appropriate). The arranging of different ggplot2 plots into panels was facilitated by the patchwork R package.[51] Smoothed density plots of exact (fractional) age were constructed with Gaussian kernels, and smoothing bandwidths were set equal to the standard deviations of the kernels. For various figures, outcomes automatically output in decimal degrees were transformed to meters for ease of interpretation. As Nicaragua is close to the equator, 1 decimal degree was taken to be equal to 111.32 kilometers for conversion purposes.

Predicted outcomes on the absolute scale were visualized with color palettes of the viridis and viridisLite R packages.[52,53] All analysis operations were performed on the continuous output of spatial models. However, mapped continuous data were discretized during the data visualization step at increments of five percentage points to better interpret color-coded outcomes at specific locations. All scales bars in the main figures were dynamic such that they only contained the colors of the corresponding panel, even if the range of the scale bars were larger. When using the vis.gam() function of the mgcv R package, it was not possible to clip the resulting contour plot to the polygon of the study area. Thus, analyses that relied on this function for output retained the default continuous color output and overlaid contour lines to better demarcate intervals on the prediction surface.

### Supplementary Tables

**Table S1.** Distribution of age among the PDCS populations, by epidemic.

| | ChikE1 | ChikE2 | ZikaE |
|---|---|---|---|
| **Female (%)** | 1,571 (50.3%) | 1,441 (50.3%) | 1,516 (50.2%) |
| **0-1 years old (%)** | 0 (0.0%) | 0 (0.0%) | 0 (0.0%) |
| **2-5 years old (%)** | 886 (28.4%) | 714 (24.9%) | 703 (23.3%) |
| **6-9 years old (%)** | 1,038 (33.2%) | 1,057 (36.9%) | 1,092 (36.2%) |
| **10-14 years old (%)** | 1,200 (38.4%) | 1,093 (38.2%) | 1,222 (40.5%) |

Abbreviations: ChikE1, first chikungunya epidemic; ChikE2, second chikungunya epidemics; PDCS, Pediatric Dengue Cohort Study; ZikaE, Zika epidemic

**Table S2.** SaTScan output for the purely spatial cluster analyses.[1]

| Epidemic | Cluster type, case-control comparison[2] | Cluster number[3] | Radius (km) | Cluster type | N cases[2] in cluster / overall N in cluster | SMR[4] | Relative risk[5] | p-value |
|---|---|---|---|---|---|---|---|---|
| ChikE1 | Infection incidence, infected participants (cases) vs. uninfected participants (controls) | 1 | 0.003 (a single house) | Gini | 5 / 5 | 15.78 | 16.16 | 0.003 |
| | | 2 | 0.78 | Hierarchical | 63 / 1517 | 0.66 | 0.49 | 0.014 |
| | | 3 | 0.35 | Gini | 3 / 296 | 0.16 | 0.15 | 0.016 |
| ChikE1[6] | Disease incidence, symptomatic infections (cases) vs. uninfected persons and subclinical infections (controls) | | | | | | | |
| ChikE2 | Infection incidence, infected participants (cases) vs. uninfected participants (controls) | 1 | 0.60 | Hierarchical | 62 / 477 | 0.52 | 0.48 | < 0.001 |
| | | 2 | 0.82 | Hierarchical | 430 / 1415 | 1.23 | 1.57 | < 0.001 |
| | | 3 | 0.29 | Gini | 70 / 155 | 1.82 | 1.91 | < 0.001 |
| | | 4 | 0.25 | Gini | 13 / 142 | 0.37 | 0.36 | 0.023 |
| ChikE2 | Disease incidence, symptomatic infections (cases) vs uninfected persons and subclinical infections (controls) | 1 | 0.81 | Hierarchical | 40 / 576 | 0.48 | 0.42 | < 0.001 |
| | | 2 | 0.28 | Gini | 47 / 141 | 2.29 | 2.46 | < 0.001 |
| | | 3 | 0.28 | Gini | 49 / 174 | 1.94 | 2.06 | 0.034 |
| | | 4 | 0.55 | Gini | 17 / 284 | 0.41 | 0.39 | 0.039 |
| ZikaE | Infection incidence, infected participants (cases) vs. uninfected participants (controls) | 1 | 1.07 | Hierarchical | 456 / 1169 | 0.83 | 0.75 | < 0.001 |
| | | 2 | 0.94 | Hierarchical | 629 / 1149 | 1.17 | 1.30 | < 0.001 |
| | | 3 | 0.21 | Gini | 98 / 143 | 1.46 | 1.49 | 0.003 |
| | | 4 | 0.06 | Gini | 30 / 34 | 1.88 | 1.90 | 0.009 |
| | | 5 | 0.27 | Gini | 41 / 194 | 0.59 | 0.57 | 0.016 |
| ZikaE | Disease incidence, symptomatic infections (cases) vs. uninfected persons and subclinical infections (controls) | 1 | 0.88 | Gini | 197 / 860 | 1.40 | 1.66 | < 0.001 |
| | | 2 | 1.07 | Hierarchical | 129 / 1113 | 0.71 | 0.60 | 0.002 |
| | | 3 | 0.67 | Gini | 103 / 892 | 0.71 | 0.63 | 0.042 |

[1]Only data for statistically significant clusters is presented in this table, as is the norm in spatial epidemiology.

[2]Cluster detection analyses use case-control terminology differently than this paper. Whereas we use *case* to denote a symptomatic infection, *case* means the group of interest and *control* means the control group in cluster detection nomenclature. Thus, when assessing the risk of infection, for example, infected participants constitute the case group and uninfected participants constitute the control. We apply the case-control nomenclature of cluster detection analyses to this table as that is the standard in the field of spatial health research.

[3]Cluster number is with respect to Figure 3.

[4]The SRM is a measure of association that compares the observed number of outcomes in the cluster to the expected number of outcomes in the cluster if (under the null) it had exhibited the same proportion of outcomes as the entire study area.

[5]The relative risk (risk ratio) is a measure of association that compares the proportion of outcomes in the cluster to the proportion of outcomes outside the cluster.

[6]No significant clusters for this outcome were identified.

Abbreviations: ChikE1, first chikungunya epidemic; ChikE2, second chikungunya epidemic; SMR, standardized mortality ratio; ZikaE, Zika epidemic

**Table S3.** Odds ratios and 95% CIs for infection, estimated from increasingly complex logistic models.[1,2]

| Epidemic | Variable | Non-spatial GLM | Non-spatial GLMM[3] | Spatial GLMM, without household adjustment[4] | Spatial GLMM, with household adjustment[5] |
|---|---|---|---|---|---|
| ChikE1 | Distance to cemetery (per 100m) | 0.98 (95% CI: 0.96, 1.01) | 0.98 (95% CI: 0.94, 1.02) | 0.99 (95% CI: 0.93, 1.05) | 0.99 (95% CI: 0.92, 1.05) |
| | Age (per 1 year) | **1.09 (95% CI: 1.05, 1.13)** | **1.13 (95% CI: 1.07, 1.20)** | **1.10 (95% CI: 1.05, 1.15)** | **1.10 (95% CI: 1.05, 1.15)** |
| | Female sex | 0.86 (95% CI: 0.64, 1.14) | 0.79 (95% CI: 0.54, 1.15) | 0.82 (95% CI: 0.61, 1.12) | 0.83 (95% CI: 0.61, 1.13) |
| | Hours w/o water (per 1 hour) | 1.01 (95% CI: 0.98, 1.04) | 1.03 (95% CI: 0.98, 1.08) | 1.01 (95% CI: 0.97, 1.05) | 1.01 (95% CI: 0.97, 1.05) |
| ChikE2[6] | Distance to cemetery (per 100m) | **0.96 (95% CI: 0.94, 0.97)** | **0.94 (95% CI: 0.92, 0.97)** | **0.96 (95% CI: 0.93, 0.99)** | **0.96 (95% CI: 0.93, 0.99)** |
| | Age (per 1 year) | **1.13 (95% CI: 1.10, 1.16)** | **1.17 (95% CI: 1.13, 1.21)** | **1.13 (95% CI: 1.10, 1.17)** | **1.13 (95% CI: 1.10, 1.17)** |
| | Female sex | 1.03 (95% CI: 0.86, 1.22) | 1.00 (95% CI: 0.81, 1.25) | 1.02 (95% CI: 0.84, 1.22) | 1.00 (95% CI: 0.83, 1.21) |
| | Hours w/o water (per 1 hour) | 0.99 (95% CI: 0.96, 1.01) | 0.99 (95% CI: 0.96, 1.02) | 0.99 (95% CI: 0.96, 1.02) | 0.99 (95% CI: 0.96, 1.02) |
| ZikaE | Distance to cemetery (per 100m)[7] | **0.95 (95% CI: 0.94, 0.97)** | **0.95 (95% CI: 0.93, 0.97)** | **0.96 (95% CI: 0.94, 0.97)** | **0.96 (95% CI: 0.94, 0.97)** |
| | Age (per 1 year) | **1.13 (95% CI: 1.11, 1.16)** | **1.18 (95% CI: 1.14, 1.21)** | **1.14 (95% CI: 1.11, 1.17)** | **1.14 (95% CI: 1.11, 1.17)** |
| | Female sex | **1.28 (95% CI: 1.11, 1.49)** | **1.35 (95% CI: 1.12, 1.62)** | **1.28 (95% CI: 1.10, 1.50)** | **1.29 (95% CI: 1.10, 1.51)** |
| | Hours w/o water (per 1 hour) | 1.01 (95% CI: 0.99, 1.03) | 1.02 (95% CI: 0.99, 1.05) | 1.01 (95% CI: 0.98, 1.03) | 1.01 (95% CI: 0.98, 1.03) |

[1]Statistically significant covariates are bolded. Some of the models do not return p-values, but whether covariates are statistically significant can be inferred from whether the CI includes the null value of 1.

[2]In general, results from generalized linear, mixed, geostatistical, and mixed geostatistical multivariable models were similar because infection and disease outcomes were not clustered within households (Table S5) and both outcomes were spatially correlated over small distances (Figure S19). Accounting for moderate-to-substantial levels of clustering would have resulted in enlarged CIs relative to models that do not account for clustering.

[3]These models were evaluated using Moran's I test.

[4]The spatial GLMM that does not account for household clustering is the geostatistical analogue of the non-spatial GLM.

[5]The spatial GLMM that does account for household clustering is the geostatistical analogue of the non-spatial GLMM.

[6]The non-spatial GLMM was run with the bobyqa optimizer for both optimization steps to avoid model convergence issues.

[7]Infection models for CHIKV and ZIKV were parameterized with distance to the cemetery on a 100m basis as the study area is ~3km as its widest. The seemingly small reduction in odds of infections per 100m from the cemetery compounds multiplicatively such that, during ZikaE, the conditional odds of infection among participants living 1 km from the cemetery were 0.63 (95% CI: 0.55, 0.73), as reported in the main text.

Abbreviations: ChikE1, first chikungunya epidemic; ChikE2, second chikungunya epidemic; CI, confidence interval; GLM, generalized linear model; GLMM, generalized linear mixed model

**Table S4.** Odds ratios and 95% CIs for disease, estimated from increasingly complex logistic models.[1,2]

| Epidemic | Variable | Non-spatial GLM | Non-spatial GLMM[3] | Spatial GLMM, without household adjustment[4] | Spatial GLMM, with household adjustment[5] |
|---|---|---|---|---|---|
| ChikE1 | Age (per 1 year) | **1.16 (95% CI: 1.09, 1.23)** | **1.17 (95% CI: 1.10, 1.27)** | **1.16 (95% CI: 1.09, 1.23)** | **1.16 (95% CI: 1.16, 1.23)** |
| | Female sex | 0.75 (95% CI: 0.49, 1.15) | 0.71 (95% CI: 0.43, 1.12) | 0.73 (95% CI: 0.47, 1.13) | 0.73 (95% CI: 0.47, 1.13) |
| ChikE2 | Age (per 1 year) | **1.13 (95% CI: 0.10, 1.17)** | **1.16 (95% CI: 1.12, 1.21)** | **1.13 (95% CI: 1.10, 1.17)** | **1.13 (95% CI: 1.09, 1.17)** |
| | Female sex | 0.98 (95% CI: 0.79, 1.21) | 0.95 (95% CI: 0.74, 1.22) | 0.96 (95% CI: 0.77, 1.20) | 0.96 (95% CI: 0.77, 1.19) |
| ZikaE | Age (per 1 year) | **1.09 (95% CI: 1.06, 1.13)** | **1.11 (95% CI: 1.07, 1.16)** | **1.10 (95% CI: 1.06, 1.13)** | **1.09 (95% CI: 1.06, 1.13)** |
| | Female sex | **1.30 (95% CI: 1.07, 1.58)** | **1.34 (95% CI: 1.06, 1.69)** | **1.30 (95% CI: 1.06, 1.59)** | **1.29 (95% CI: 1.05, 1.58)** |
| | Prior DENV infection | **0.79 (95% CI: 0.63, 0.99)** | **0.75 (95% CI: 0.56, 0.99)** | **0.77 (95% CI: 0.60, 0.98)** | **0.77 (95% CI: 0.61, 0.99)** |

[1]Statistically significant covariates are bolded. Some of the models do not return p-values, but whether covariates are statistically significant can be inferred from whether the CI includes the null value of 1.

[2]In general, results from generalized linear, mixed, geostatistical, and mixed geostatistical multivariable models were similar because infection and disease outcomes were not clustered within households (Table S5) and both outcomes were spatially correlated over small distances (Figure S19). Accounting for moderate-to-substantial levels of clustering would have resulted in enlarged CIs relative to models that do not account for clustering.

[3]These models were evaluated using Moran's I test.

[4]The spatial GLMM that does not account for household clustering is the geostatistical analogue of the non-spatial GLM.

[5]The spatial GLMM that does account for household clustering is the geostatistical analogue of the non-spatial GLMM.

Abbreviations: ChikE1, first chikungunya epidemic; ChikE2, second chikungunya epidemic; CI, confidence interval; DENV, dengue virus; GLM, generalized linear model; GLMM, generalized linear mixed model

**Table S5.** Intracluster correlation coefficient[1] and 95% CIs for the incidence of infection and disease within households across the three epidemics.

| Outcome of interest | Method | ChikE1 | ChikE2 | ZikaE |
|---|---|---|---|---|
| Infection incidence | ANOVA[2] | 0.22 (95% CI: 0.17, 0.28) | 0.21 (95% CI: 0.16, 0.27) | 0.22 (95% CI: 0.17, 0.27) |
| | GEE[3] | 0.24 (95% CI: 0.12, 0.35) | 0.20 (95% CI: 0.13, 0.26) | 0.18 (95% CI: 0.13, 0.23) |
| | Resampling[4] | 0.19 (95% CI: 0.00, 0.41) | 0.23 (95% CI: 0.14, 0.31) | 0.20 (95% CI: 0.13, 0.26) |
| Disease incidence[5] | ANOVA[2] | 0.04 (95% CI: 0.00, 0.09) | 0.14 (95% CI: 0.09, 0.21) | 0.16 (95% CI: 0.11, 0.21) |
| | GEE[3] | 0.08 (95% CI: 0.00, 0.17) | 0.12 (95% CI: 0.05, 0.18) | 0.13 (95% CI: 0.07, 0.19) |
| | Resampling[4] | Not estimable due to the small number of chikungunya cases | 0.17 (95% CI: 0.05, 0.29) | 0.16 (95% CI: 0.06, 0.27) |

[1]The ICC ranges from 0-1. A value of 0 indicates outcomes within homes are not correlated. A value of 1 indicates outcomes within homes are fully correlated.

[2]Point estimates were derived from the traditional one-way ANOVA-based approach, and interval estimates were derived from Searle's exact confidence limit equations.[17]

[3]Point and interval estimates were derived from a logistic GEE model using an exchangeable correlation structure.[2] GEE models for CHIKV and ZIKV infection incidence were adjusted for age, sex, water availability, and distance to the cemetery. GEE models for chikungunya incidence were adjusted for age and sex. GEE models for Zika incidence were adjusted for age, sex, and prior DENV infection.

[4]Point and interval estimates derived from the modern resampling-based approach of Chakraborty and Sen.[19]

[5]The correlation of infection outcomes within households was the primary analysis; we have no reason to believe, based on the existing literature, that disease outcomes clusters within households. However, we present estimates of the ICC for disease outcomes to compare with the ICC values for infection outcomes.

Abbreviations: ANOVA, analysis of variance; ChikE1, first chikungunya epidemic; ChikE2, second chikungunya epidemic; CHIKV, chikungunya virus; CI, confidence interval; GEE, generalized estimating equations; ICC, intracluster correlation coefficient; ZIKV, Zika virus; ZikaE, Zika epidemic

### Supplemental Figures

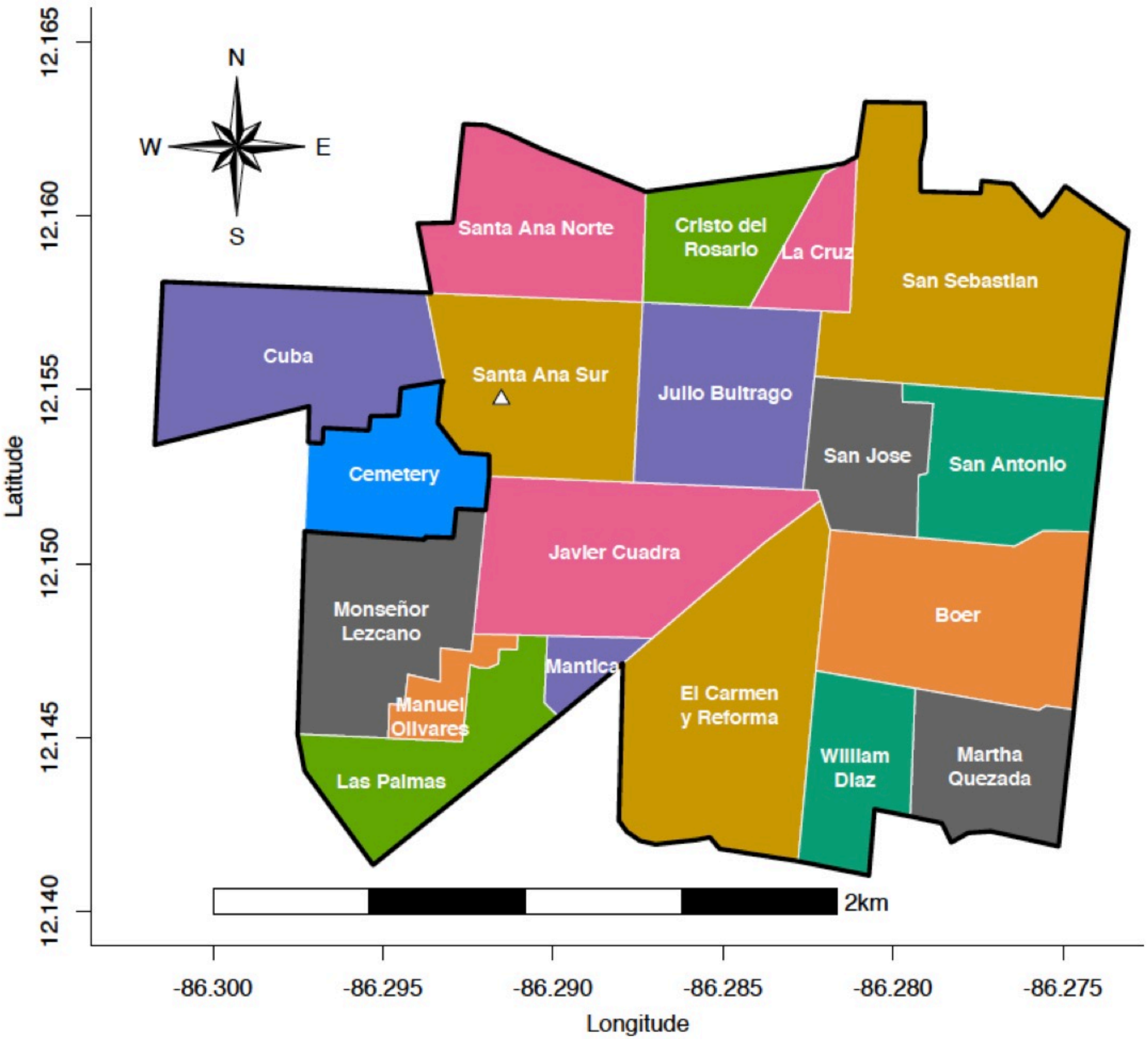


**Figure S1.** The neighborhoods of the study area in Managua, Nicaragua. The cemetery is shown in blue, and the study health center is indicated by a white triangle.

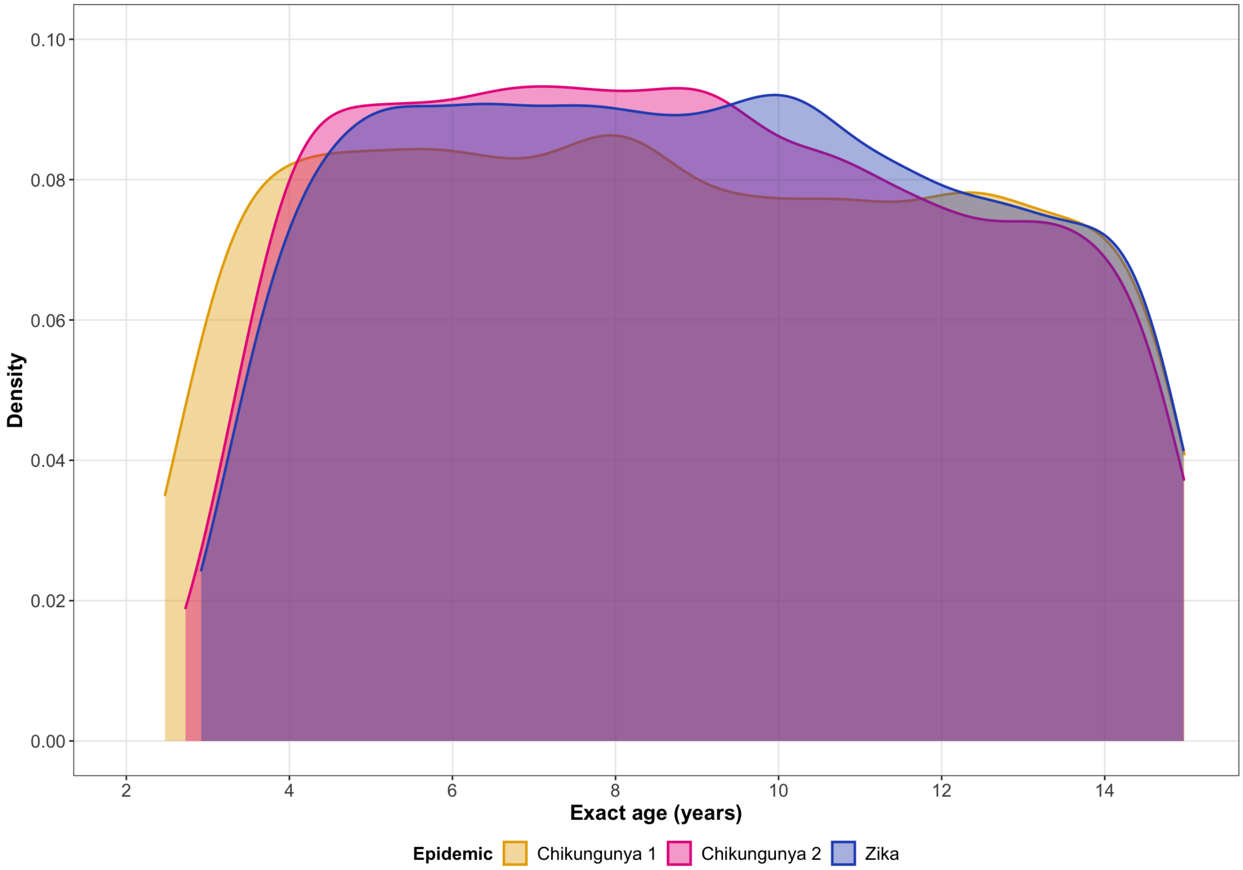


**Figure S2.** Distribution of exact age among PDCS participants by epidemic period.

Abbreviations: PDCS, Pediatric Dengue Cohort Study

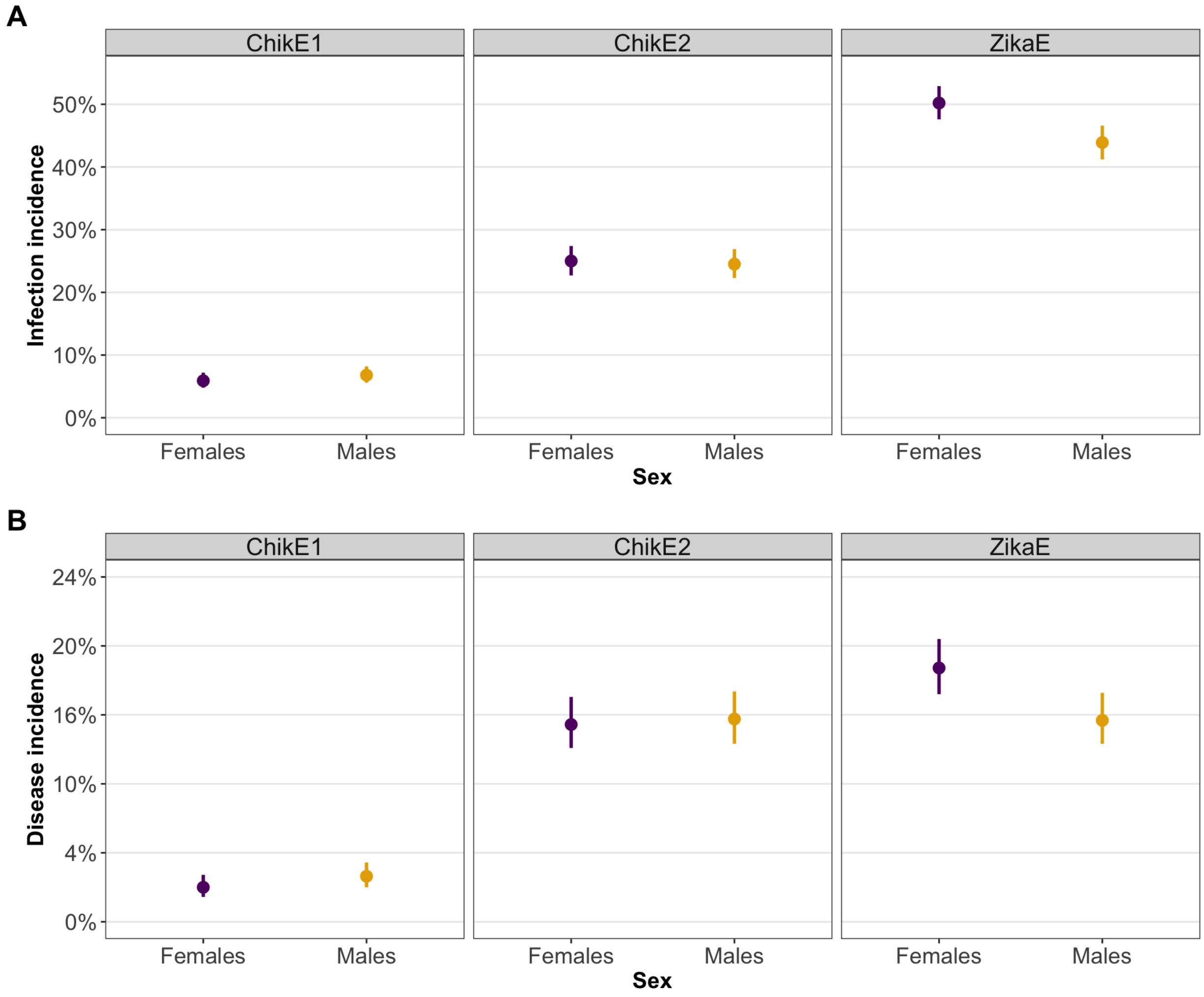


**Figure S3**. Point and interval (95% confidence interval) estimates for the infection incidence **(A)** and the disease incidence (**B**) by epidemic and sex. There was a significant risk difference (difference in proportions) for infection incidence during ZikaE; female children had a higher incidence of ZIKV infection (risk difference = 6.1%; 95% CI: 2.6, 9.5; p-value < 0.001) than male children. There was a significant risk difference for the disease incidence during ZikaE; females had a higher incidence of Zika (risk difference = 3.7%; 95% CI: 1.1, 6.2; p-value = 0.004) than males. All other comparisons were not significant.

Abbreviations: ChikE1, first chikungunya epidemic; ChikE2, second chikungunya epidemic; CI, confidence interval; ZikaE, Zika epidemic; ZIKV, Zika virus

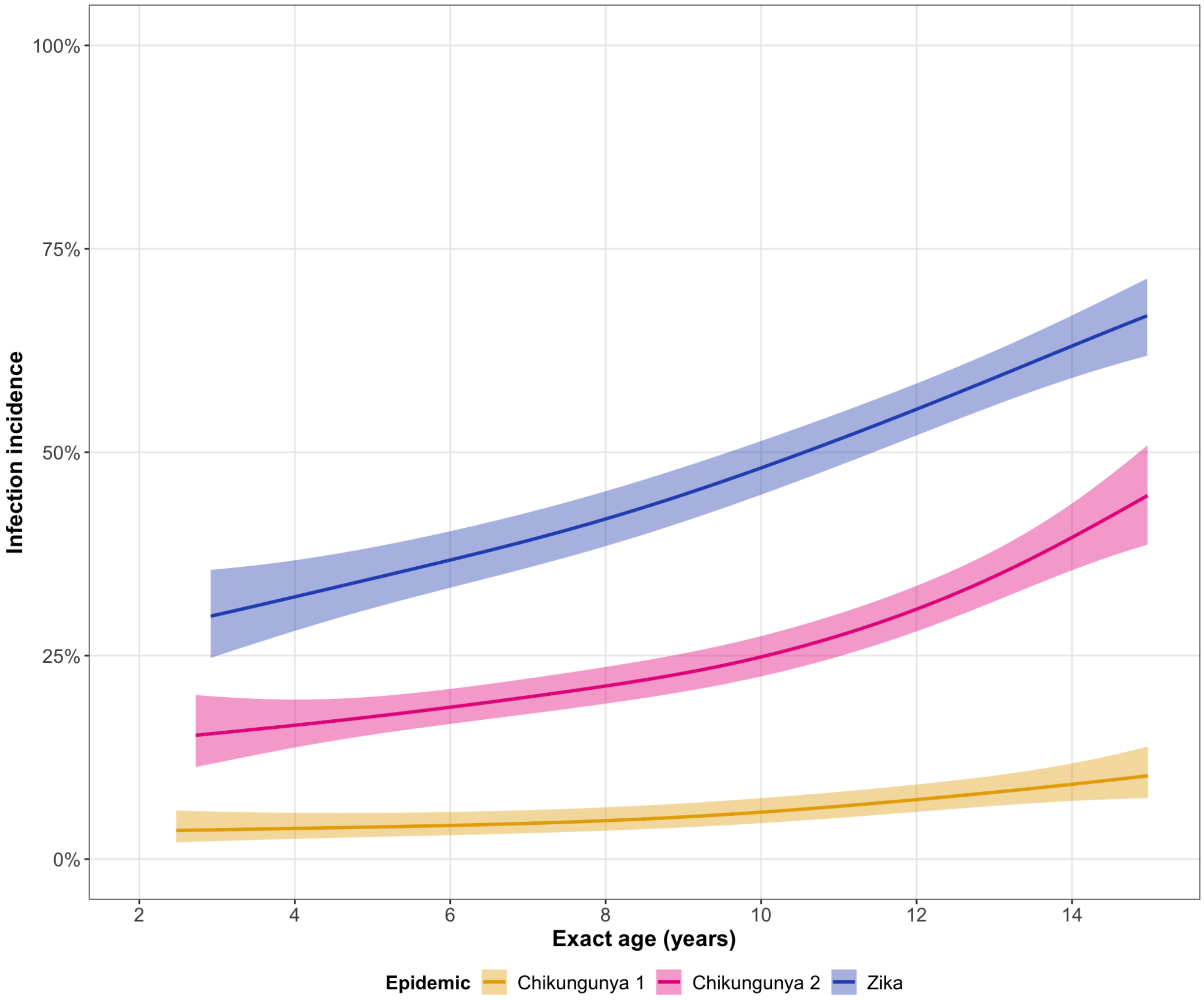


**Figure S4.** Age trends for the infection incidence, along with pointwise 95% confidence bands, for the first chikungunya epidemic, the second chikungunya epidemic, and the Zika epidemic across the age range of the PDCS (2-14 years of age).

Abbreviations: PDCS, Pediatric Dengue Cohort Study

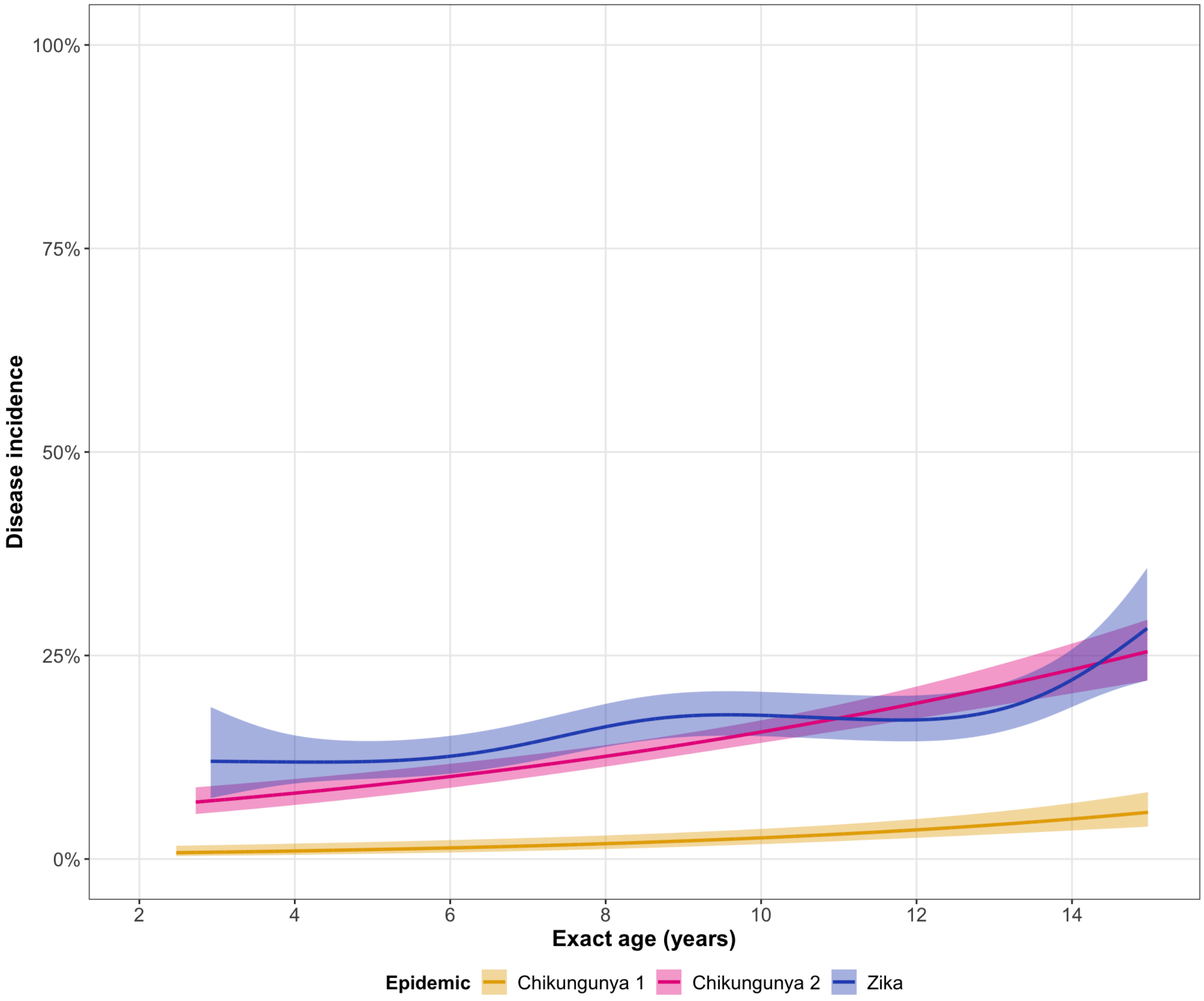


**Figure S5.** Age trends for the disease incidence, along with pointwise 95% confidence bands, for the first chikungunya epidemic, the second chikungunya epidemic, and the Zika epidemic across the age range of the PDCS (2-14 years of age).

Abbreviations: PDCS, Pediatric Dengue Cohort Study

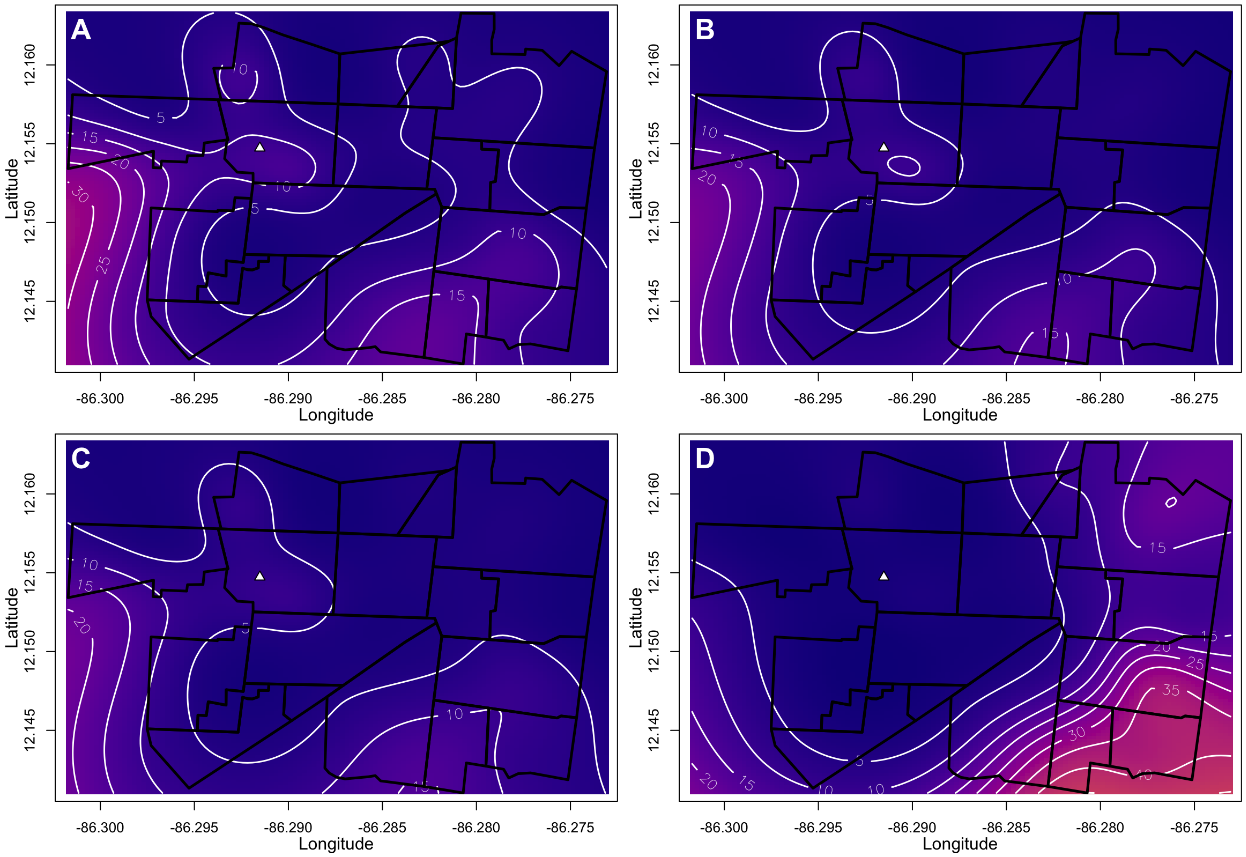


**Figure S6.** CHIKV infection incidence estimated from spatial logistic GAMs during the first chikungunya epidemic. Panels show **(A)** unadjusted infection incidence; **(B)** infection incidence adjusted for age and sex; **(C)** infection incidence adjusted for age, sex, and water availability; and **(D)** infection incidence adjusted for age, sex, water availability, and distance to the cemetery. The variables in models for Panels B-D have been set to those of the median participant across the three PDCS epidemics, a female of age 8.71 years living in a household that has 24-hour indoor access to tap water and is located 841.50 meters from the boundary of the local cemetery. Figures S6-8 and S12-14 use the same color scale and median values to facilitate cross-epidemic comparisons.

Abbreviations: CHIKV, chikungunya virus; GAM, generalized additive model; PDCS, Pediatric Dengue Cohort Study

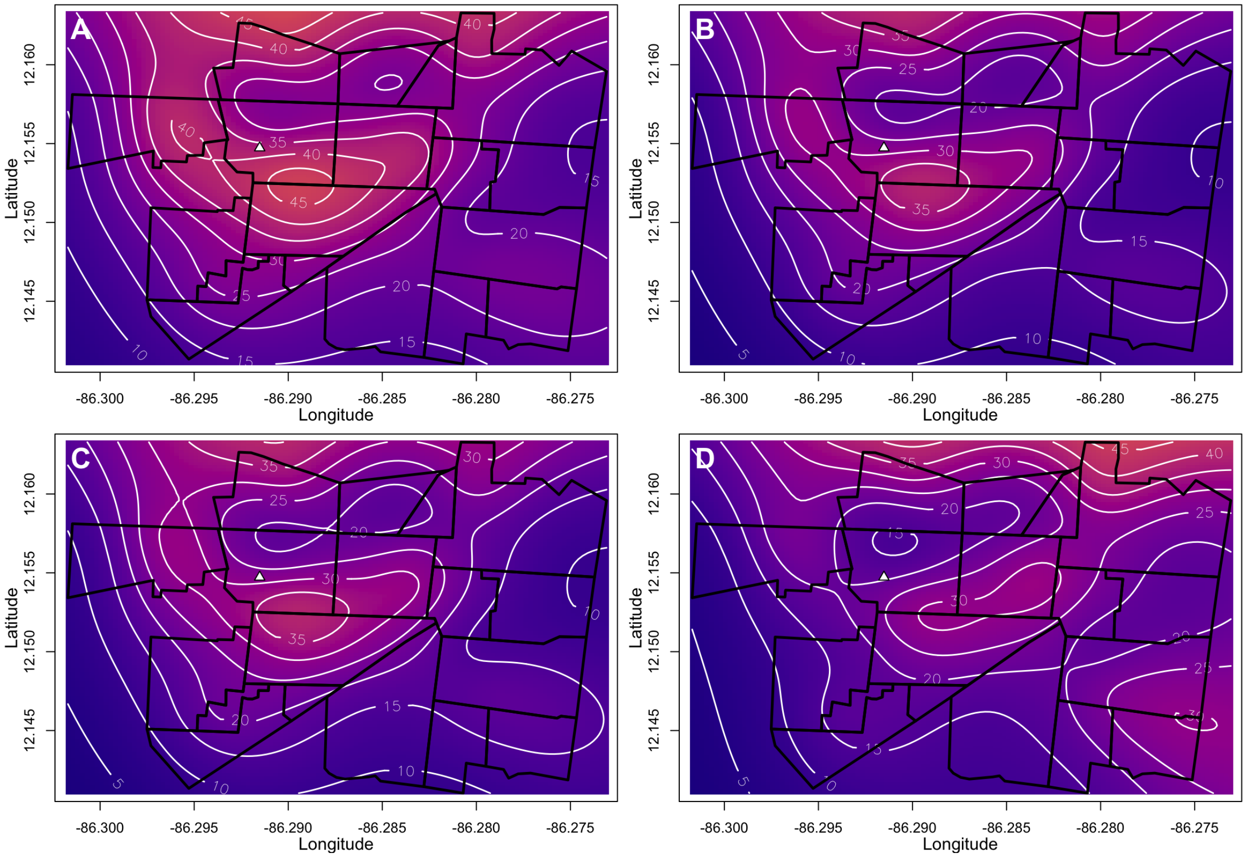


**Figure S7.** CHIKV infection incidence estimated from spatial logistic GAMs during the second chikungunya epidemic. Panels show **(A)** unadjusted incidence; **(B)** infection incidence adjusted for age and sex; **(C)** infection incidence adjusted for age, sex, and water availability; and **(D)** infection incidence adjusted for age, sex, water availability, and distance to the cemetery. The variables in models for Panels B-D have been set to those of the median participant across the three PDCS epidemics, a female of age 8.71 years living in a household that has 24-hour indoor access to tap water and is located 841.50 meters from the boundary of the local cemetery. Figures S6-8 and S12-14 use the same color scale and median values to facilitate cross-epidemic comparisons.

Abbreviations: CHIKV, chikungunya virus; GAM, generalized additive model; PDCS, Pediatric Dengue Cohort Study

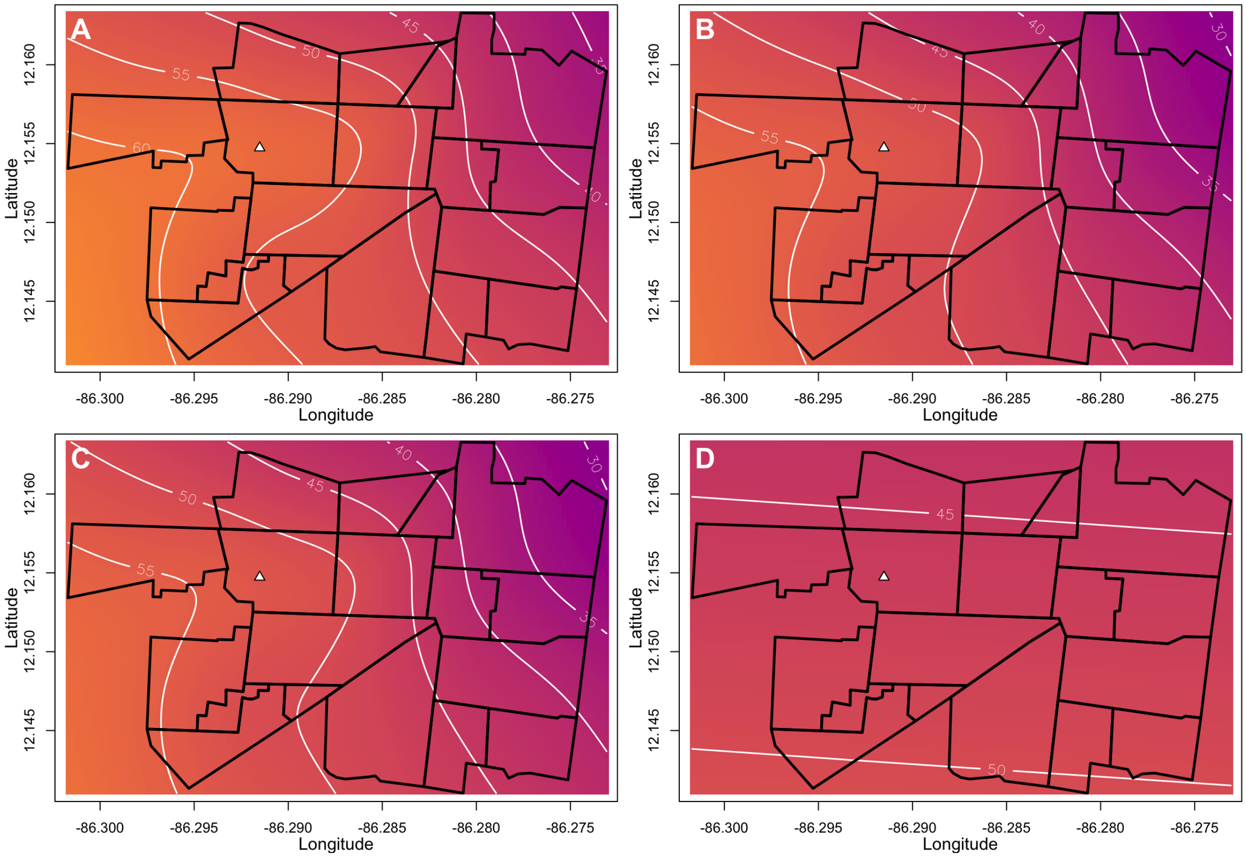


**Figure S8.** ZIKV infection incidence estimated from spatial logistic GAMs during the Zika epidemic. Panels show **(A)** unadjusted infection incidence; **(B)** infection incidence adjusted for age and sex; **(C)** infection incidence adjusted for age, sex, and water availability; and **(D)** infection incidence adjusted for age, sex, water availability, and distance to the cemetery. The variables in models for Panels B-D have been set to those of the median participant across the three PDCS epidemics, a female of age 8.71 years living in a household that has 24-hour indoor access to tap water and is located 841.50 meters from the boundary of the local cemetery. Figures S6-8 and S12-14 use the same color scale and median values to facilitate cross-epidemic comparisons.

Abbreviations: GAM, generalized additive model; PDCS, Pediatric Dengue Cohort Study; ZIKV, Zika virus

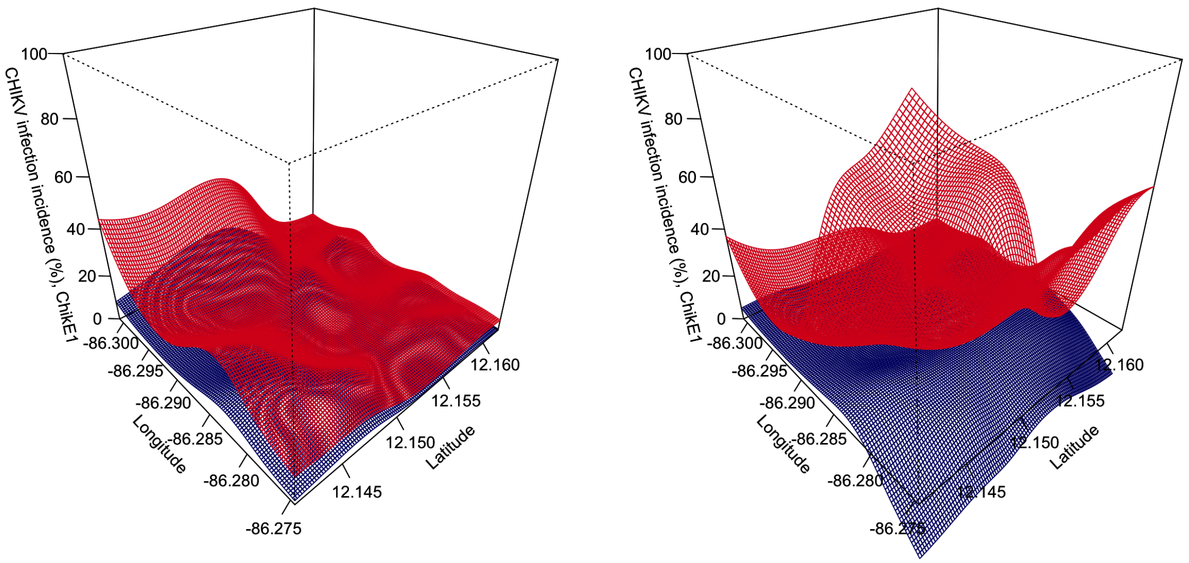


**Figure S9.** Uncertainty bounds for the unadjusted **(A)** and adjusted **(B)** (for age, sex, water availability, and distance to the cemetery) spatial incidence of infection estimates from the first chikungunya epidemic. The unadjusted and fully adjusted panels from Figure S6A,D lie between the red (point estimate + 1SE surface) and the blue (point estimate – 1SE surface). The surfaces should be interpreted as indicative of where there is more certainty (red and blue surfaces are close to each other) and where there is less certainty (red and blue surfaces are far from each other). Unlike the point estimate surfaces of Figure S6, these uncertainty surfaces are not model-constrained to be between 0-1. As a result, the surfaces may exceed the [0,1] probability space, which is why they should not be interpreted as the 3D equivalent of confidence intervals.

Abbreviations: SE, standard error

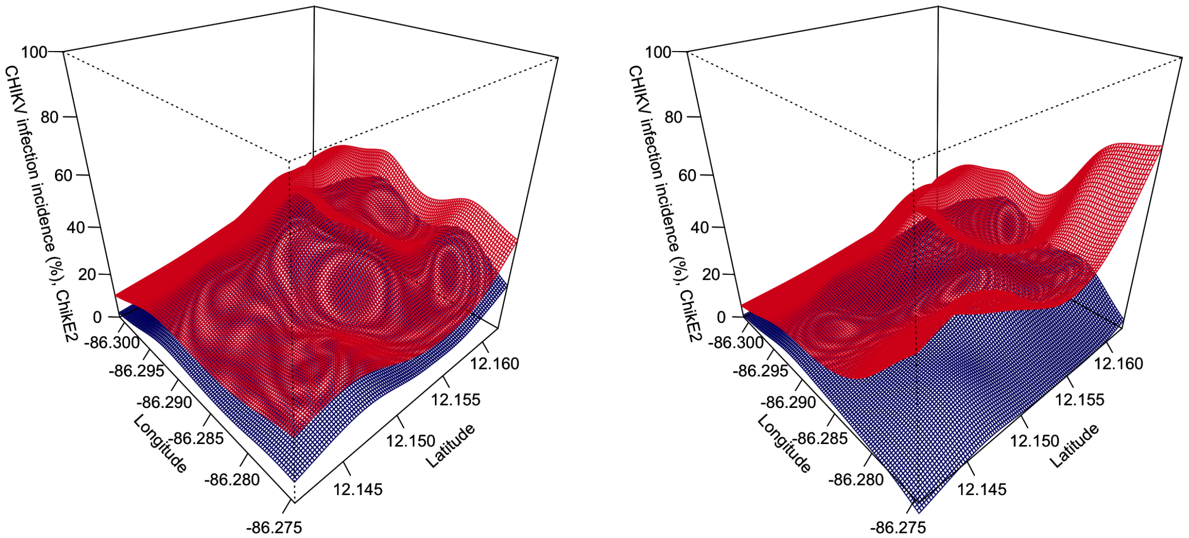


**Figure S10.** Uncertainty bounds for the unadjusted **(A)** and adjusted **(B)** (for age, sex, water availability, and distance to the cemetery) spatial incidence of infection estimates from the second chikungunya epidemic. The unadjusted and fully adjusted panels from Figure S7A,D lie between the red (point estimate + 1SE surface) and the blue (point estimate – 1SE surface). The surfaces should be interpreted as indicative of where there is more certainty (red and blue surfaces are close to each other) and where there is less certainty (red and blue surfaces are far from each other). Unlike the point estimate surfaces of Figure S7, these uncertainty surfaces are not model-constrained to be between 0-1. As a result, the surfaces may exceed the [0,1] probability space, which is why they should not be interpreted as the 3D equivalent of confidence intervals.

Abbreviations: SE, standard error

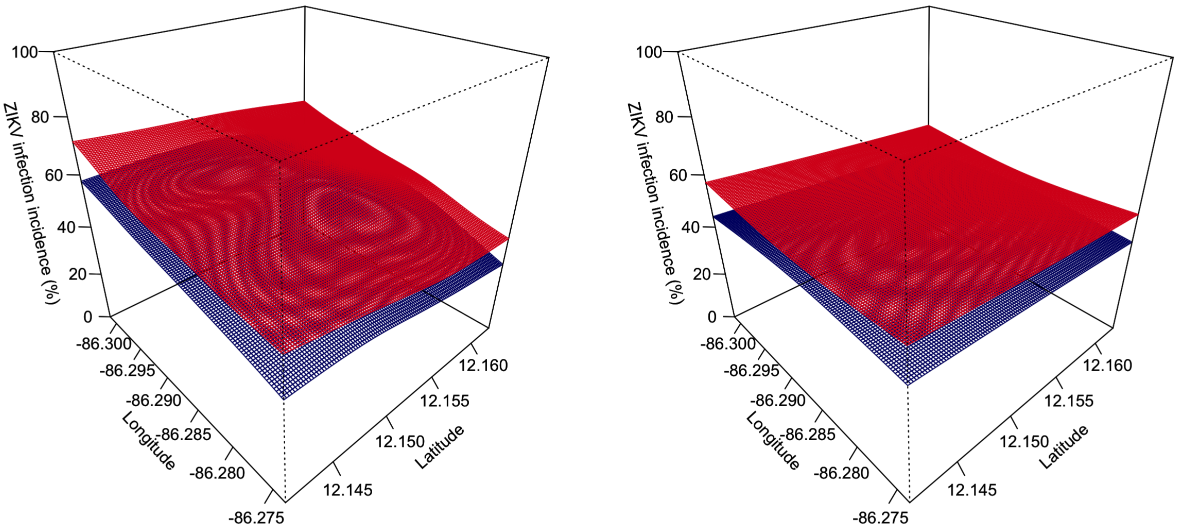


**Figure S11.** Uncertainty bounds for the unadjusted **(A)** and adjusted **(B)** (for age, sex, water availability, and distance to the cemetery) spatial incidence of infection estimates from the Zika epidemic. The unadjusted and fully adjusted panels from Figure S8A,D lie between the red (point estimate + 1SE surface) and the blue (point estimate – 1SE surface). The surfaces should be interpreted as indicative of where there is more certainty (red and blue surfaces are close to each other) and where there is less certainty (red and blue surfaces are far from each other). Unlike the point estimate surfaces of Figure S8, these uncertainty surfaces are not model-constrained to be between 0-1. As a result, the surfaces may exceed the [0,1] probability space, which is why they should not be interpreted as the 3D equivalent of confidence intervals

Abbreviations: SE, standard error

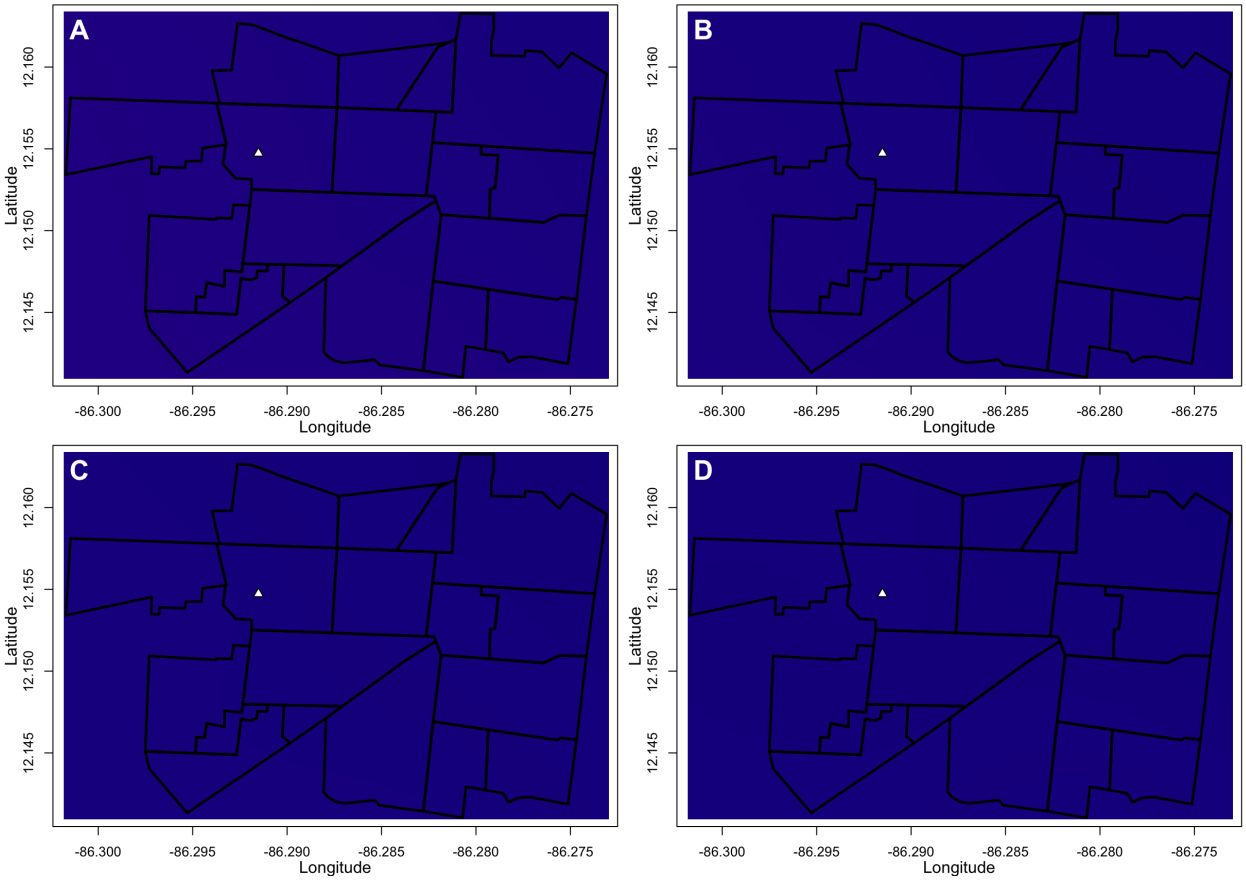


**Figure S12.** Disease incidence estimated from spatial logistic GAMs during the first chikungunya epidemic. Panels show **(A)** unadjusted disease incidence; **(B)** disease incidence adjusted for age; **(C)** disease incidence adjusted for age and sex; and **(D)** disease incidence adjusted for age, sex, and distance to the cemetery. The variables in models for Panels B-D have been set to those of the median participant across the three PDCS epidemics, a female of age 8.71 years living in a household that has 24-hour indoor access to tap water and is located 841.50 meters from the boundary of the local cemetery. Disease incidence was so low during the first chikungunya epidemic that all mapped values are between 0-5%. Figures S6-8 and S12-14 use the same color scale and median values to facilitate cross-epidemic comparisons.

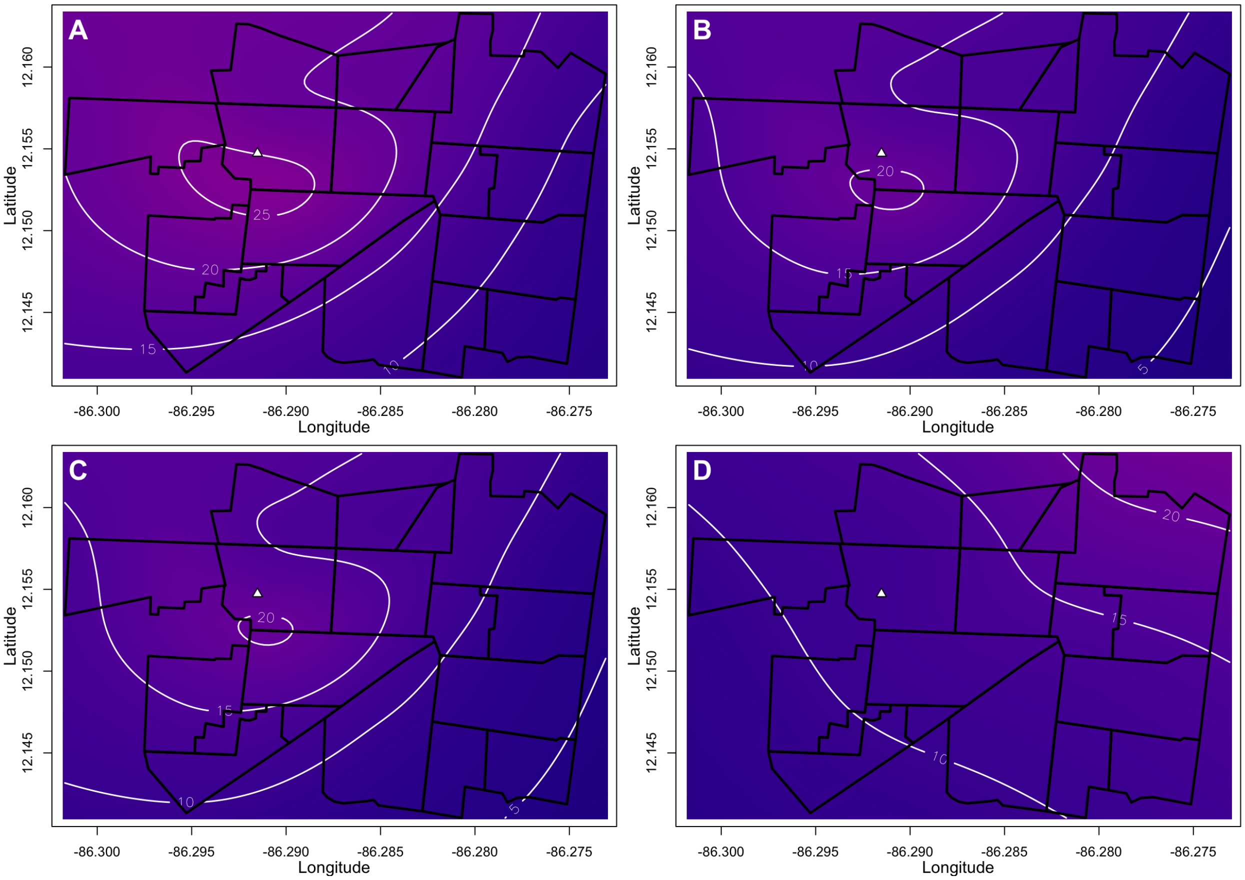


**Figure S13.** Disease incidence estimated from spatial logistic GAMs during the second chikungunya epidemic. Panels show **(A)** unadjusted disease incidence; **(B)** disease incidence adjusted for age; **(C)** disease incidence adjusted for age and sex; and **(D)** disease incidence adjusted for age, sex, and distance to the cemetery. The variables in models for Panels B-D have been set to those of the median participant across the three PDCS epidemics, a female of age 8.71 years living in a household that has 24-hour indoor access to tap water and is located 841.50 meters from the boundary of the local cemetery. Figures S6-8 and S12-14 use the same color scale and median values to facilitate cross-epidemic comparisons.

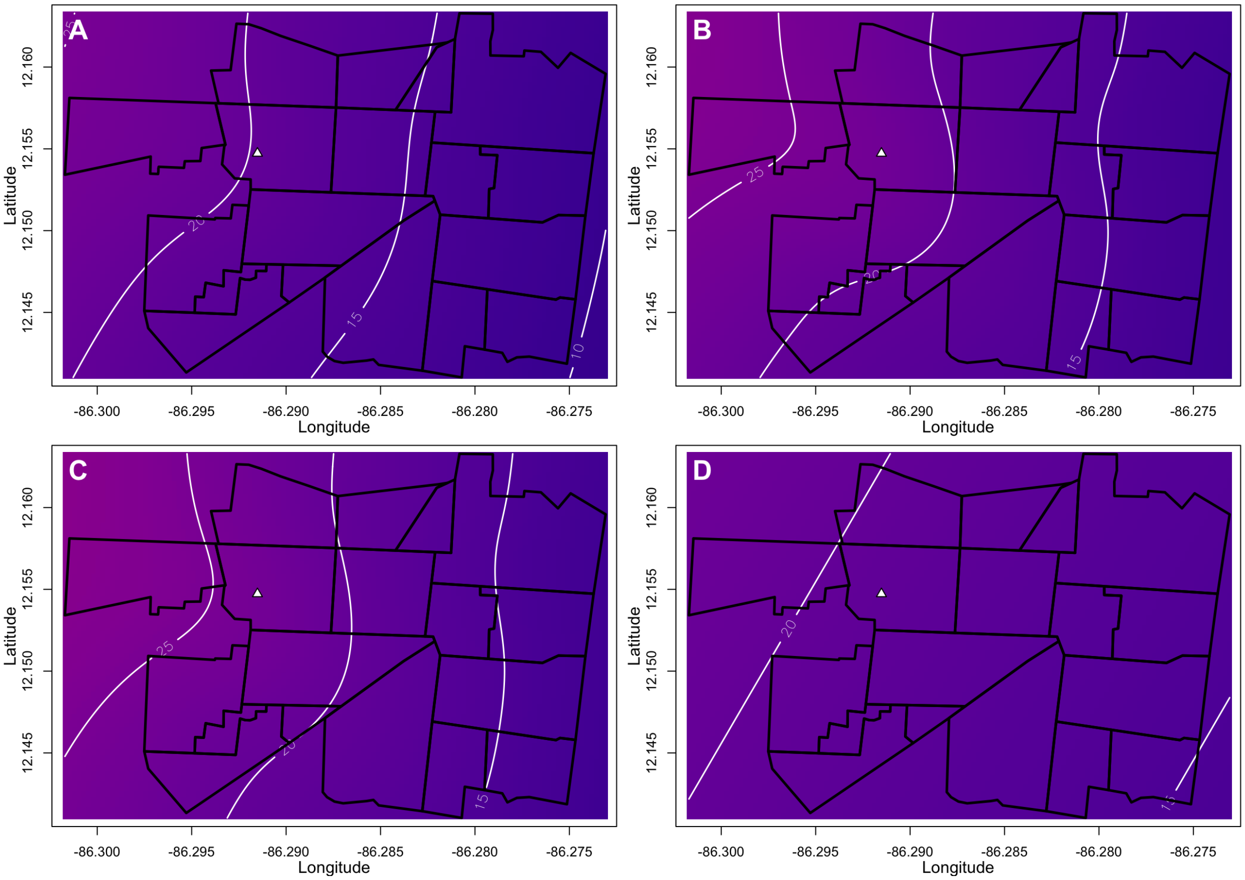


**Figure S14.** Disease incidence estimated from spatial logistic GAMs during the Zika epidemic. Panels show **(A)** unadjusted disease incidence; **(B)** disease incidence adjusted for age and sex; **(C)** disease incidence adjusted for age, sex, and prior dengue virus infection status; and **(D)** disease incidence adjusted for age, sex, prior dengue virus infection status, and distance to the cemetery. The variables in models for Panels B-D have been set to those of the median participant across the three PDCS epidemics, a female of age 8.71 years living in a household that has 24-hour indoor access to tap water and is located 841.50 meters from the boundary of the local cemetery. Figures S6-8 and S12-14 use the same color scale and median values to facilitate cross-epidemic comparisons.

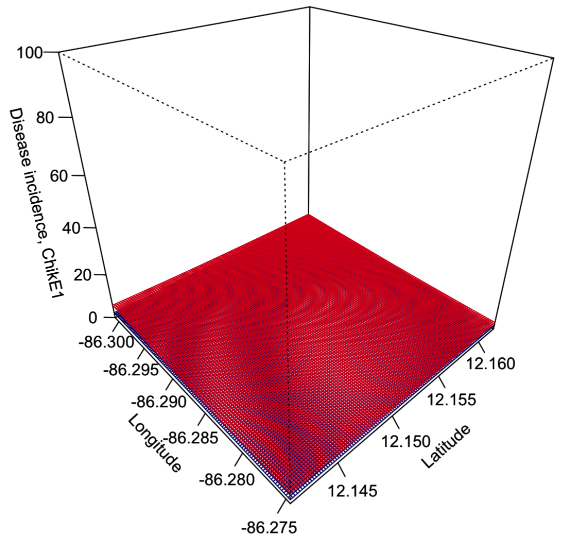


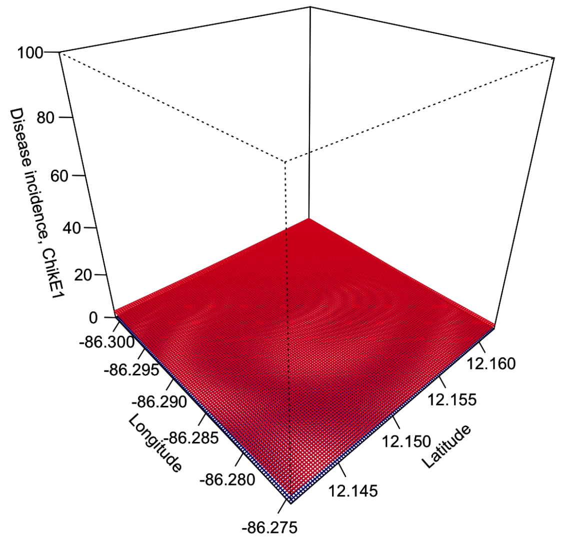


**Figure S15.** Uncertainty bounds for the unadjusted **(A)** and adjusted **(B)** (for age, sex, and distance to the cemetery) spatial incidence of disease estimates from the first chikungunya epidemic. The unadjusted and fully adjusted panels from Figure S12A,D lie between the red (point estimate + 1SE surface) and the blue (point estimate – 1SE surface). The surfaces should be interpreted as indicative of where there is more certainty (red and blue surfaces are close to each other) and where there is less certainty (red and blue surfaces are far from each other). Unlike the point estimate surfaces of Figure S12, these uncertainty surfaces are not model-constrained to be between 0-1. As a result, the surfaces may exceed the [0,1] probability space, which is why they should not be interpreted as the 3D equivalent of confidence intervals.

Abbreviations: SE, standard error

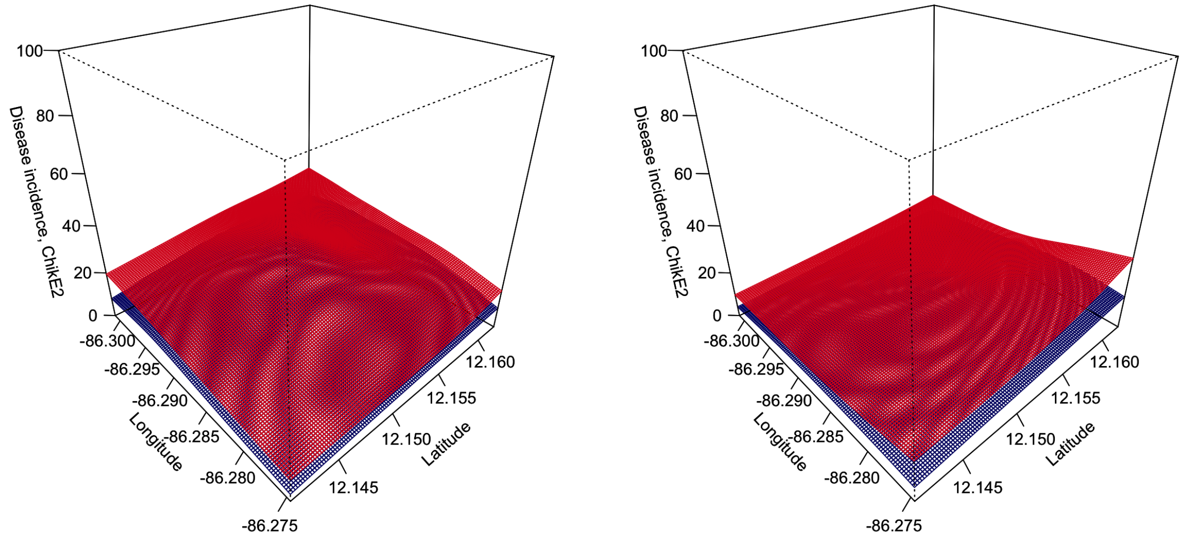


**Figure S16.** Uncertainty bounds for the unadjusted **(A)** and adjusted **(B)** (for age, sex, and distance to the cemetery) spatial incidence of disease estimates from the second chikungunya epidemic. The unadjusted and fully adjusted panels from Figure S13A,D lie between the red (point estimate + 1SE surface) and the blue (point estimate − 1SE surface). The surfaces should be interpreted as indicative of where there is more certainty (red and blue surfaces are close to each other) and where there is less certainty (red and blue surfaces are far from each other). Unlike the point estimate surfaces of Figure S13, these uncertainty surfaces are not model-constrained to be between 0-1. As a result, the surfaces may exceed the [0,1] probability space, which is why they should not be interpreted as the 3D equivalent of confidence intervals.

Abbreviations: SE, standard error

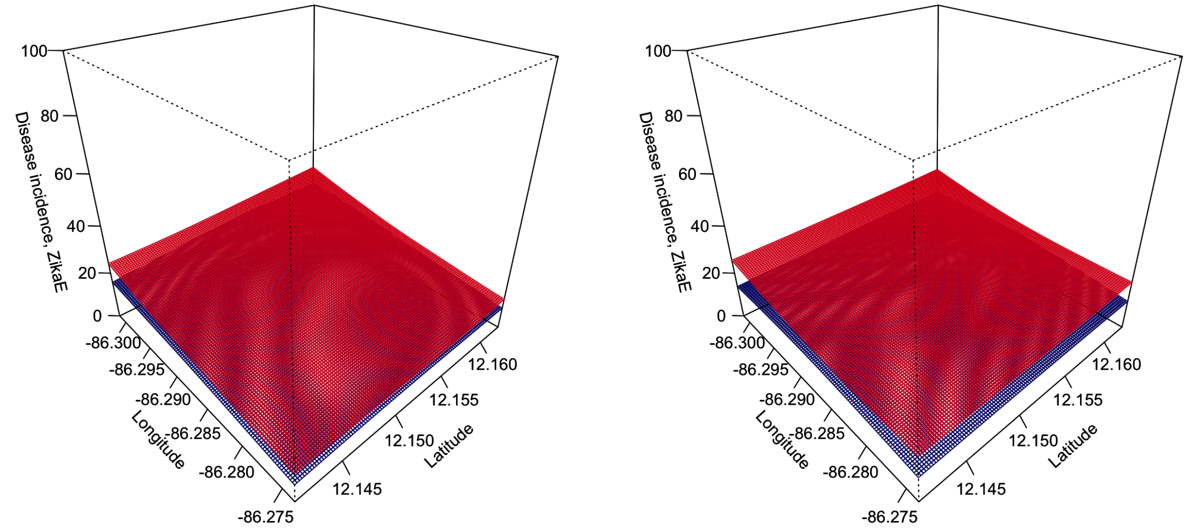


**Figure S17.** Uncertainty bounds for the unadjusted **(A)** and adjusted **(B)** (for age, sex, prior dengue virus infection status, and distance to the cemetery) spatial incidence of disease estimates from the Zika epidemic. The unadjusted and fully adjusted panels from Figure S14A,D lie between the red (point estimate + 1SE surface) and the blue (point estimate – 1SE surface). The surfaces should be interpreted as indicative of where there is more certainty (red and blue surfaces are close to each other) and where there is less certainty (red and blue surfaces are far from each other). Unlike the point estimate surfaces of Figure S14, these uncertainty surfaces are not model-constrained to be between 0-1. As a result, the surfaces may exceed the [0,1] probability space, which is why they should not be interpreted as the 3D equivalent of confidence intervals.

Abbreviations: SE, standard error

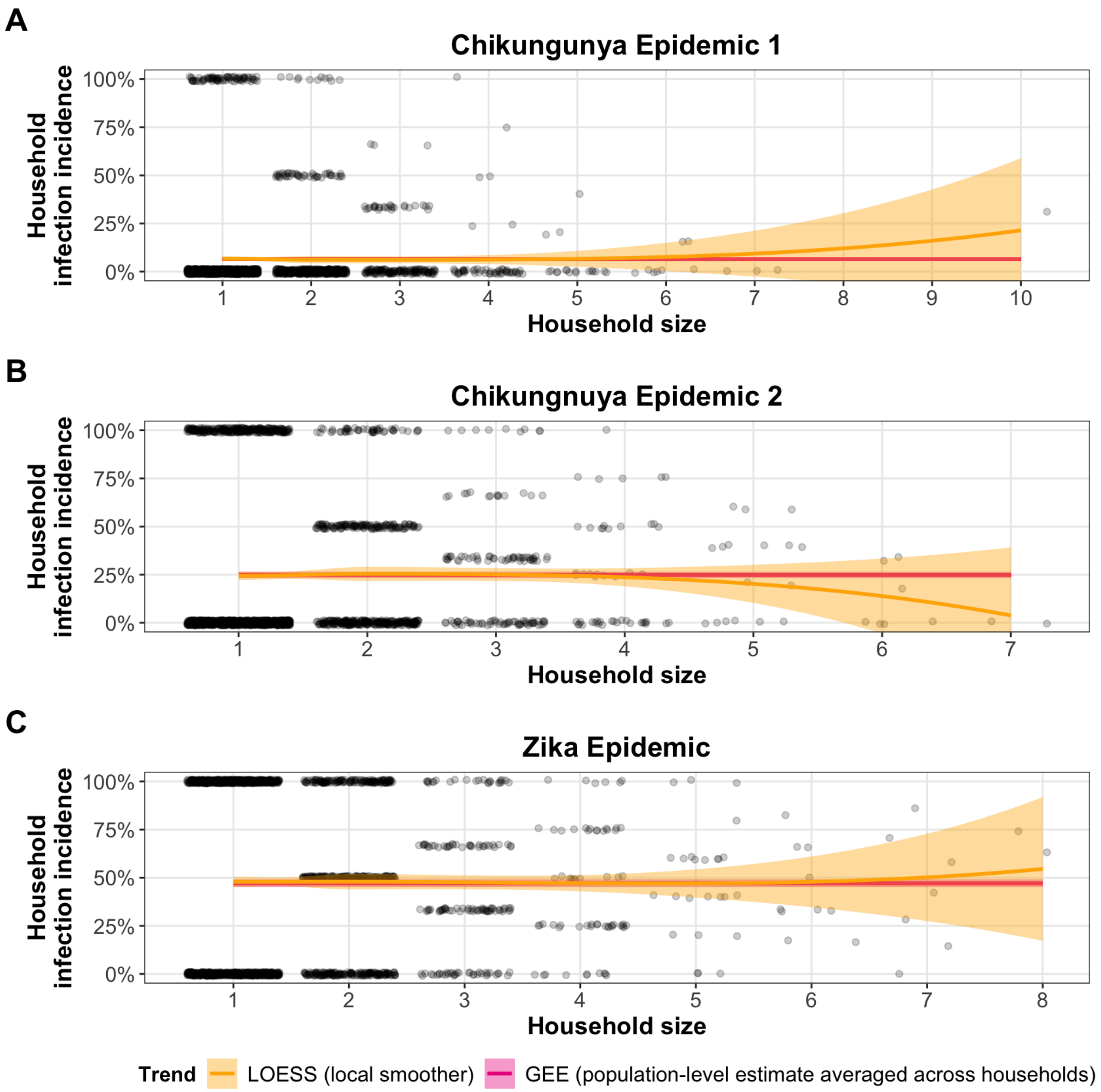


**Figure S18.** The association between household size and household infection incidence (number of household members infected / household size) during **(A)** ChikE1, **(B)** ChikE2, and **(C)** ZikaE. Where there were sufficient data to reliably estimate a trend (PDCS households with ≤5 participants), there was no evidence of household infection incidence scaling with household size. Both the independent and dependent variables for this analysis refer only to PDCS participants, as we have no infection data on household members who are not PDCS participants. The yellow line is a smooth LOESS trend. The pink line is the population-averaged, intercept-only GEE estimate of infection incidence, which averages over households of different sizes. A 95% confidence band is shown for each estimate in the corresponding color. Note that the confidence band for the LOESS trend dips below 0 because there is no numerical constraint on the LOESS smoother. A GAM approach failed to converge because of insufficient variation in values of household infection incidence, which prompted the use of the less robust LOESS algorithm.

Abbreviations: ChikE1, chikungunya epidemic 2; ChikE2, chikungunya epidemic 2; GEE, generalized estimating equations; LOESS, locally estimated scatterplot smoothing; GAM, generalized additive model; ZikaE, Zika epidemic

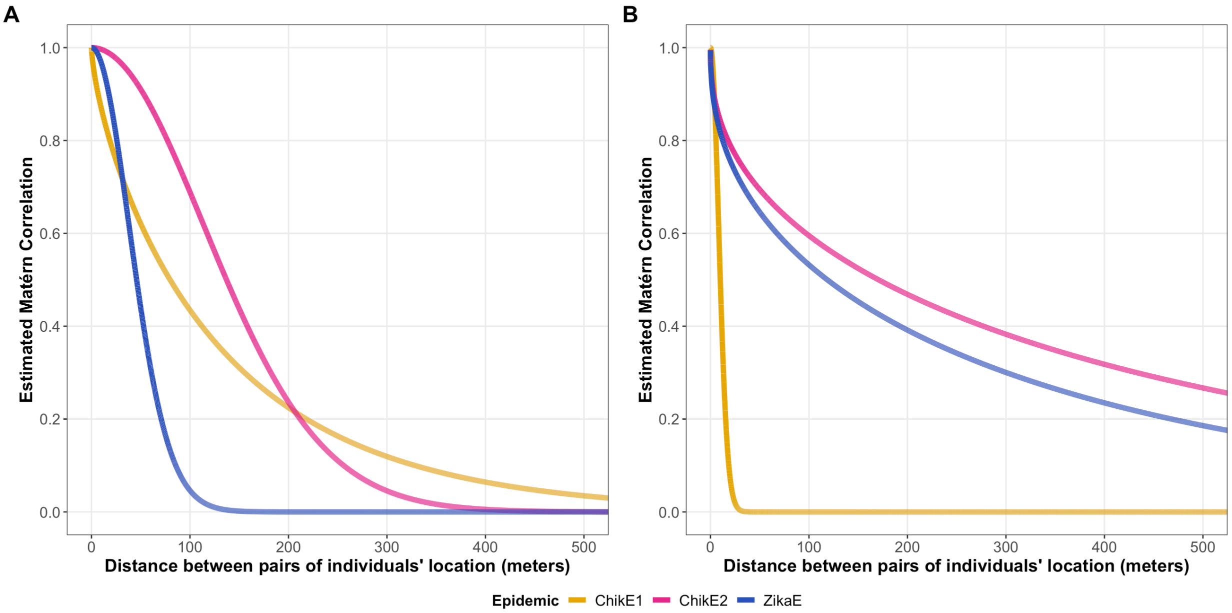


**Figure S19.** The spatial correlation of infection outcomes **(A)** and disease outcomes **(B)** across the three epidemics we considered. Panel A was the primary analysis. The analysis for disease outcomes among all participants, Panel B, is shown for completeness. The estimated Matérn autocorrelation function for infection and disease outcomes between two locations at varying distances is shown. For example, the CHIKV infection outcomes of persons living >200 meters apart during ChikE1 and ChikE2 have a correlation <0.2, on average. The Matérn autocorrelation parameters underlying the functions shown were estimated from the logistic geostatistical mixed models for infection and disease that account for spatial and household correlation (rightmost columns of Tables S3-S4). As the disease status (regardless of CHIKV or ZIKV infection) for one person is biologically unlikely to affect the disease status of another (also regardless of infection), the slow decay of the autocorrelation function for disease outcomes during ChikE2 and ZikaE suggests that unknown variables relevant to the spatial distribution of chikungunya and Zika occurrence were not included in their respective models.

Abbreviations: ChikE1, chikungunya epidemic 1; ChikE2, chikungunya epidemic 2; CHIKV, chikungunya virus; ZikaE, Zika epidemic; ZIKV, Zika virus

45. Vyas S, Kumaranayake L. Constructing socio-economic status indices: how to use principal components

analysis. Health Policy Plan. 2006 Aug 30;21(6):459–68.

46. Sturrock H. Fitting point process models to disease case data with mgcv (or whatever you want). GitHub. 2020. Available from: https://github.com/disarm-platform/disarmr/blob/master/RMarkdowns/ppm_mgcv.md

47. Pebesma E. Simple features for R: Standardized support for spatial vector data. R J. 2018;10(1):439–46.

48. Sturrock H. disarmr: A collection of spatial analysis functions related to the DiSARM project at UCSF. 2020.

49. Wickham H. ggplot2: Elegant Graphics for Data Analysis. Springer-Verlag New York; 2016.

50. Wickham H. ggplot2. 1st ed. New York City: Springer; 2009. 1–212 p.

51. Pedersen TL. patchwork: The Composer of Plots. 2019.

52. Garnier S. viridis: Default Color Maps from "matplotlib." 2018.

53. Garnier S, Ross N, Rudis R, Camargo A, Sciaini M, Scherer C. viridis - colorblind-friendly color maps for R. 2021.
